# Mechanisms linking physical activity with psychiatric symptoms across lifespan: A systematic review

**DOI:** 10.1101/2022.07.13.22277479

**Authors:** Phuong Thuy Nguyen Ho, Tram Ha Pham Bich, Thao Tong, Wichor M Bramer, Amy Hofman, David Revalds Lubans, Meike W. Vernooij, María Rodriguez-Ayllon

## Abstract

**Background:** Physical activity has been suggested as a protective factor against psychiatric symptoms. While numerous studies have focused on the magnitude of physical activity’s effect on psychiatric symptoms, few have examined the potential mechanisms.

**Objective:** The current review aimed to synthesize scientific evidence of the mechanisms through which physical activity might reduce psychiatric symptoms across the lifespan.

**Methods:** We included articles that were published before March 2022 from five electronic databases (Medline, Web of Science, PsycINFO, Embase, and Cochrane). A qualitative synthesis of studies was conducted. The risk of bias assessment was performed using The Joanna Briggs Institute Critical Appraisal Tool for Systematic Reviews. Studies were included if they explored the possible mechanisms through which physical activity links with psychiatric symptoms (i.e., internalizing and externalizing symptoms) across the lifespan.

**Results:** A total of 24 articles were included (three RCTs, four non-RCTs, four prospective longitudinal studies, and 13 cross-sectional studies). Overall, most of the studies (70%) focused on children, adolescents, and young adults. Our findings show that self-esteem, self-concept, and self-efficacy were the only consistent paths through which physical activity may influence psychiatric symptoms (specifically depressive and anxiety symptoms). There were insufficient studies to determine the role of neurobiological and behavioral mechanisms. Findings from our systematic review suggest that self-esteem, self-concept, and self-efficacy are potential paths through which physical activity might reduce psychiatric symptoms across the lifespan.

**Conclusions:** Overall, future physical activity interventions with the purpose of improving mental health should consider these mechanisms to develop more effective interventions. Current literature gaps and recommendations for researchers to explore other potential mechanisms were also included.

**Protocol Registration:** The protocol of this study was registered in the PROSPERO database (registration number CRD42021239440) and published in April 2022.

**Key Points:**

- Self-esteem, self-concept, and self-efficacy are potential paths through which physical activity might reduce psychiatric symptoms (specifically depressive and anxiety symptoms) across the lifespan. Future studies should consider incorporating strategies to enhance these psychosocial mechanisms in physical activity interventions.
- Few studies have examined the role of neurobiological and behavioral mechanisms. It is recommended that future research could focus on these areas.
- Integrated studies that examine the combined and independent contributions of the neurobiological, psychosocial and behavioral mechanisms are needed to obtain the overall picture.
- There is a lack of research on externalizing and other internalizing symptoms beside depressive and anxiety symptoms (e.g., somatic symptoms).

## 1. Introduction

According to Lancet Global Health (2020), approximately one billion people around the world are suffering from at least one mental disorder [1]. The economic loss that came as a result of this was estimated to be $2.5 trillion per year in 2010 and could potentially increase up to $6 trillion per year in 2030. It is, therefore, not surprising that mental health problems have an enormous impact on many aspects of our lives. While researchers and experts are looking into solutions to these problems, there is still an urgent need for better understanding of mental disorders and effective preventions.

Previous studies have shown that most mental health disorders have emerged in approximately 50% of individuals by the age of 18 [2]. Mental health disorders can be classified into externalising and internalising problems. Externalizing problems include disinhibited/externally-focused behavioral symptoms such as conduct problems, rule-breaking behavior, attention-deficit/hyperactivity problems [3]. Internalizing symptoms include over-inhibited/internally-focused symptoms, such as depression, anxiety, and somatic symptoms. Having externalizing and internalizing symptoms in childhood and adolescents can predict mental illnesses later in life [4]. Even though researchers have found many risk factors for internalizing and externalizing symptoms, less is known about their protective factors [5,6].

A growing body of research has identified physical activity as a potential protective factor against psychiatric symptoms and disorders [7]. Physical activity is defined as “any bodily movement produced by skeletal muscle that requires energy expenditure” [8] and has been widely studied due to its extensive health-related benefits [9]. For instance, previous research has demonstrated that physical activity has a small-to-moderate effect on mental health; however, the underlying mechanisms responsible for these effects are unclear [10–13]. Understanding the mechanisms linking physical activity with psychiatric symptoms will allow for better explanation and prediction, and for more effective interventions. This could stimulate the identification of cost-efficient alternative therapies for preventing mental illnesses of all ages.

In 2016, Lubans et al. proposed a conceptual model with three groups of mechanisms (i.e., neurobiological, psychosocial, and behavioral mechanisms) that may explain the effects of physical activity on mental health in children and adolescents [14]. They also performed a systematic review and identified a lack of available evidence [15]. However, their review only included intervention studies, and although this type of design can provide evidence for cause and effect, observational studies can also provide complementary information, particularly when there is a lack of experimental evidence. In adults, only narrative reviews [14–16] have explored the mechanisms linking physical activity with psychiatric symptoms. In particular, Stillman et al. [9] suggested that physical activity might reduce internalizing symptoms via psychosocial pathways such as mood. Additionally, Kandola et al. [17] proposed some biological (e.g., neuroplasticity, inflammation, or oxidative stress) and psychosocial (e.g., self-esteem or social support) mechanisms underlying the relationship between physical activity and depressive symptoms in adults. Nevertheless, no previous systematic review has synthesized the existing evidence in adults.

We aim to fill in the current literature gap by synthesizing the current findings and updating all relevant literature mapping the mechanisms through which physical activity might reduce psychiatric symptoms across the lifespan. In particular, we investigated the three broad categories of mechanisms (psychosocial, neurobiological, and behavioral) as suggested by Lubans et al. [14]. Additionally, we included both intervention and observational studies and we did not impose an age limit to allow for better generalization of our findings. The findings of our review may guide future research to develop more effective treatments and solutions to protect people against mental disorders.

## 2. Methods

Our reviewed adhered to the Preferred Reporting Items for Systematic Review and Meta-Analysis (PRISMA) guidelines [18]. The design of the present work was fully specified in advance. It was registered in the PROSPERO database with the registration number CRD42021239440. Further details on the protocol can be found elsewhere [19]. It was made publicly available before conducting the primary electronic search [19].

### 2.1 Search Strategy and Inclusion Criteria

We searched five electronic databases for relevant articles: Medline All via Ovid, Web of Science Core Collection, PsycINFO via Ovid, Embase via Embase.com, and Cochrane CENTRAL via Wiley (see Online Supplemental Appendix for the full search strategies). Articles included for screening were published before March 2022 (date last searched). In brief, we included articles based on predefined criteria as summarized in the protocol [19]. Two reviewers (PTNH and THPB) screened the titles and abstracts of the articles separately and included articles based on the inclusion criteria [19]. Any disagreements were discussed and resolved between the two reviewers. Final decisions were made by a third reviewer (MR-A). The eligible articles were retrieved in full-text and screened again by the same reviewers to determine full eligibility.

The complete methodology, procedures and inclusion/exclusion criteria have been previously described [19]. Briefly, the inclusion criteria were: (1) including at least one group of healthy participants (i.e., no diagnosis of neuropsychogical disorders); (2) studies that explore the possible mechanisms through which physical activity links with psychiatric symptoms; (3) the outcome variable belongs to a psychiatric symptom, specifically an internalizing (i.e., depression, anxiety, somatic symptoms) or an externalizing (i.e., conduct problems, rule-breaking behavior, attention deficit/hyperactivity problems) symptom; (4) intervention studies (RCT, non-RCT) and observational studies (prospective longitudinal cohort studies, cross-sectional studies). An additional search for studies was performed by screening reference lists of included studies and their citations through Google Scholar. Lastly, we contacted an expert (DL) in the field to identify additional studies that may have been missed and any relevant ongoing or unpublished studies.

We did not include conference abstracts and other types of grey literature [20]. Studies that included professional athletes, animals, or only included participants with neuropsychological and/or physical disorders were excluded. Interestingly, our preliminary search showed that the majority of existing literature included behavioral mechanisms as independent variables rather than as potential mechanisms, which complicated the search and added more than 10,000 irrelevant articles. Therefore, to identify relevant articles for the behavioral mechanisms, we screened reference lists of the articles that we included in the current study and contacted an expert (DL) in the field.

### 2.2 Data extraction

Two researchers (PTNH and TT) extracted the data. This process was double-checked by one experienced researcher (MR-A). The from the eligible articles possible disagreements that occurred with the extraction were discussed by the researchers until a consensus was reached.

We used a standardized tool to obtain study background (title, author, year, country), sample characteristics (sample size, mean age, percentage of female participants), design (intervention [RCT or non-RCT], or observational [cross-sectional or longitudinal]), independent variables, dependent variables, mediating variables, instruments used to assess the variables, statistical analyses and software, confounders, and main findings. For intervention studies (RCTs and non-RCTs), the time length of intervention, description of the program, intensity, duration, and frequency were also extracted. For longitudinal studies, we also extracted years of follow-up.

### 2.3 Risk of bias

The risk of bias of each study was evaluated independently by two researchers (PTNH and TT). Any disagreements were resolved in a consensus meeting with the third researcher (MR-A). The Joanna Briggs Institute Critical Appraisal Tool for Systematic Reviews (https://jbi.global/critical-appraisal-tools) was used to assess the risk of bias. The Joanna Briggs Institute Critical Appraisal Tool for Systematic Reviews includes a checklist with specific criteria to assess the risk of bias for each study design. For each criterion, there are four possible options: “yes” (criterion met), “no” (criterion not met), “unclear” or “not applicable”. There are eight criteria items for cross-sectional studies, eleven criteria items for longitudinal (or cohort) studies, nine criteria items for non-RCTs (or quasi-experimental studies) and thirteen criteria items for RCTs. To classify the risk of bias, we used the method that was previously employed by Molina-Garcia et al. [21]. If at least 75% of the applicable items had been scored as “yes”, then the study was labeled as “low risk”. If less than 75% of the applicable items had been scored as “no”, then the study was as “high risk”. Any criteria item that was “not applicable” was included in the calculation of the percentage.

### 2.4 Data Synthesis

Findings were synthesized using a method that was previously employed by Sallis et al. [13,14,22]. If 0–33% of studies reported a statistically significant path (e.g., self-esteem) through which physical activity affect or be associated with any psychiatric symptoms, the result was classified as no association (Ø); if 34–59% of studies, or if fewer than four studies reported a significant path, the result was classified as being inconsistent/uncertain (?). Finally, if ≥ 60% of studies found a statistically significant path, the result was classified as significant (✓).

### 2.5 Modifications to the Initial Protocol

In April 2022, we published the protocol for the current study which outlined our plan to carry out the systematic review in details [19]. We originally planned to employ the Grading of Recommendations Assessment, Development and Evaluation (GRADE) framework to assess the quality of the evidence across studies. One of the criteria of the GRADE framework is publication bias, which would require us to perform a meta-analysis [23]. Due to the heterogeneity of the studies, we were unable to carry out a meta-analysis of the results, and hence, couldn’t employ the GRADE framework.

## 3. RESULTS

### 3.1 Selection process

The original search of five databases yielded 12,239 articles, 7,647 of which were identified as duplicates and removed before screening. There were 4,592 remaining articles to screen for titles and abstracts where 4,424 articles did not meet the inclusion criteria. This left us with 168 articles for the full-text screening. Out of 168 articles, only four articles were unretrievable. There were 149 articles which did not meet the inclusion criteria based on full-text screening (more details in **Table S1**). Five articles were included after screening the references of the studies that met the inclusion criteria. Finally, 24 articles were included in the review: Three RCT, four non-RCT, four prospective longitudinal studies, and 13 cross-sectional studies. Further details about the selection process are shown in **Figure 1**.

**Figure 1.**
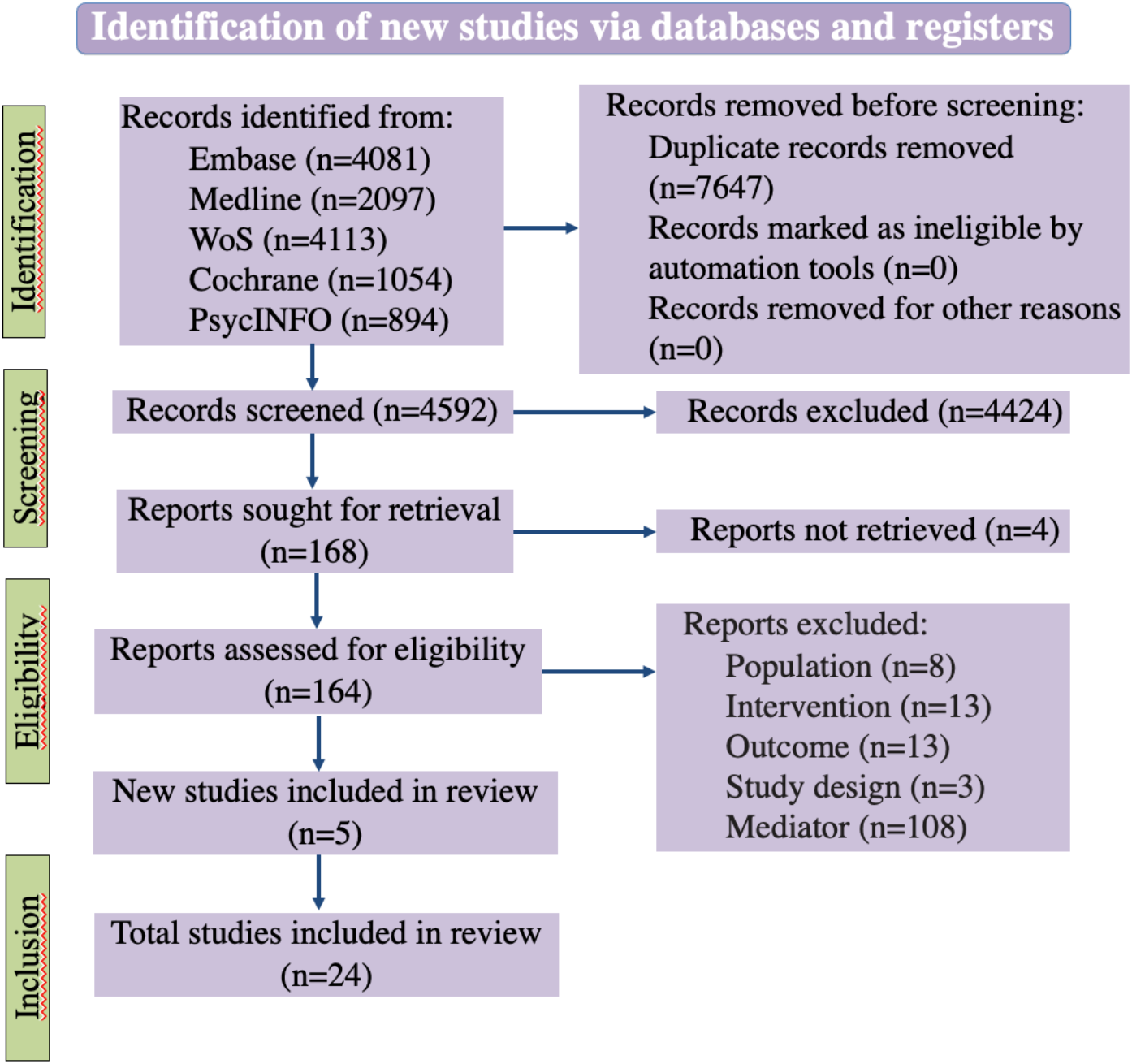
Flow diagram for study selection.

### 3.2 Summary of included studies

A detailed description of the studies included in the systematic review is provided in **Table 1**.

**Table 1.**
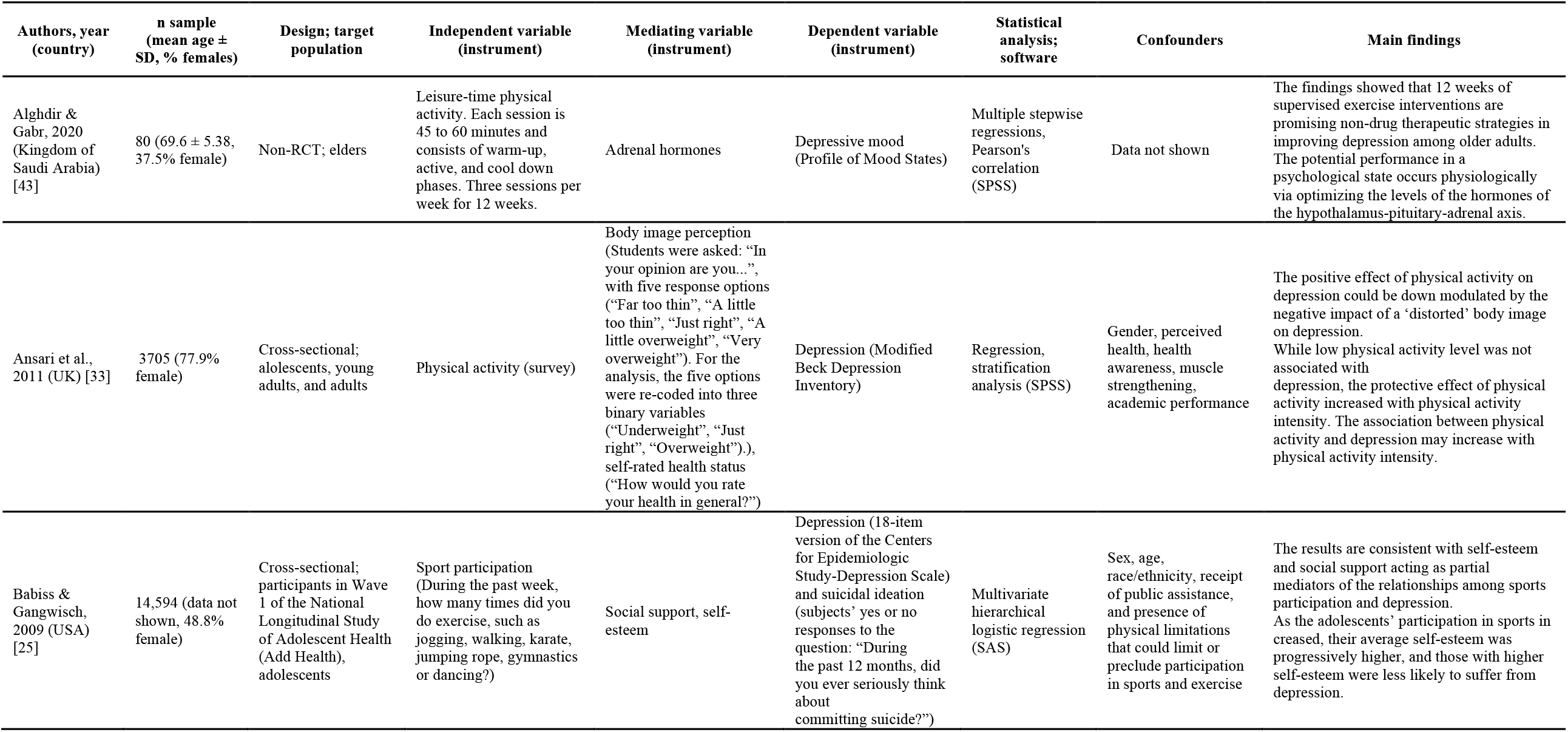
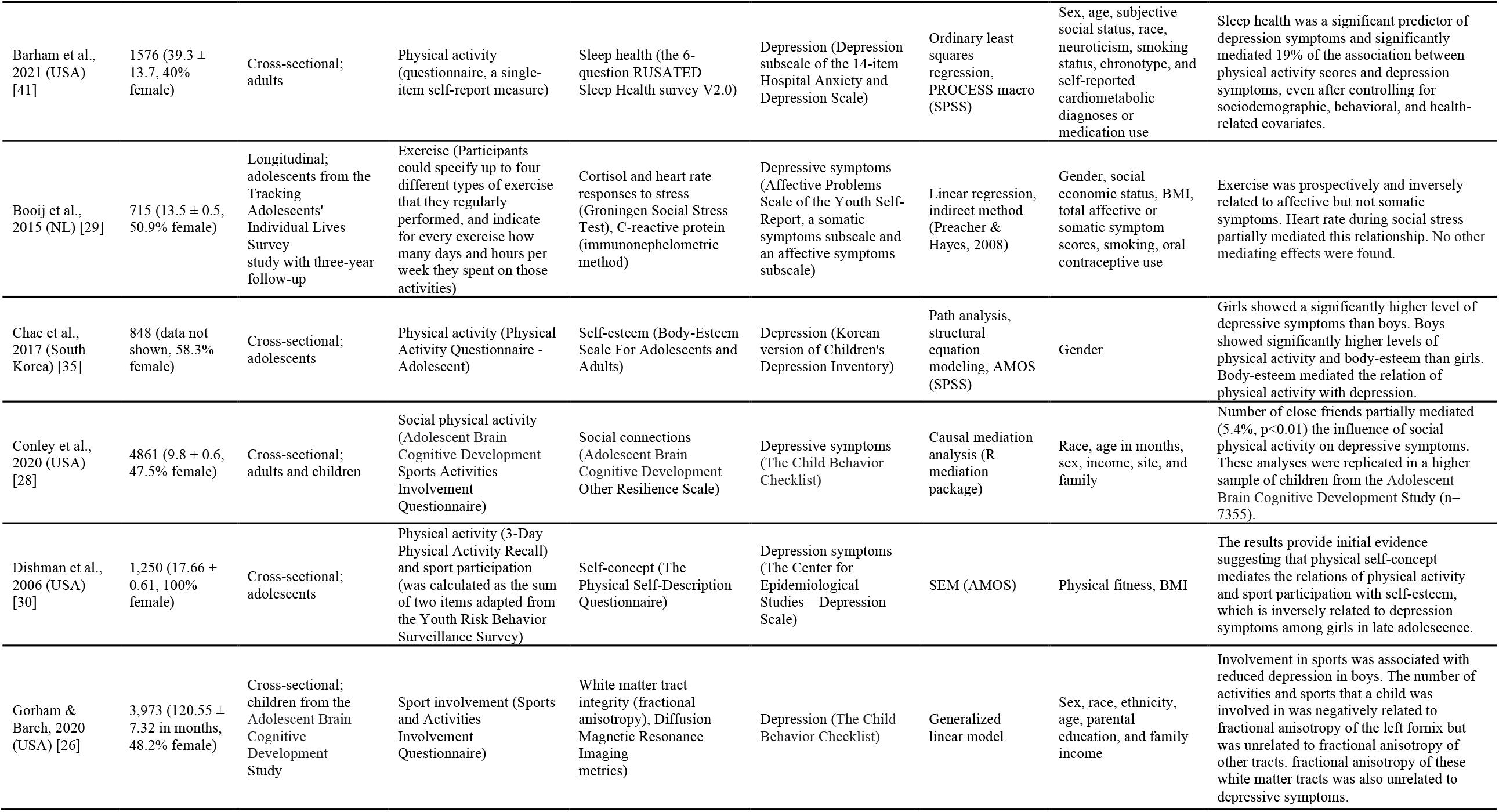
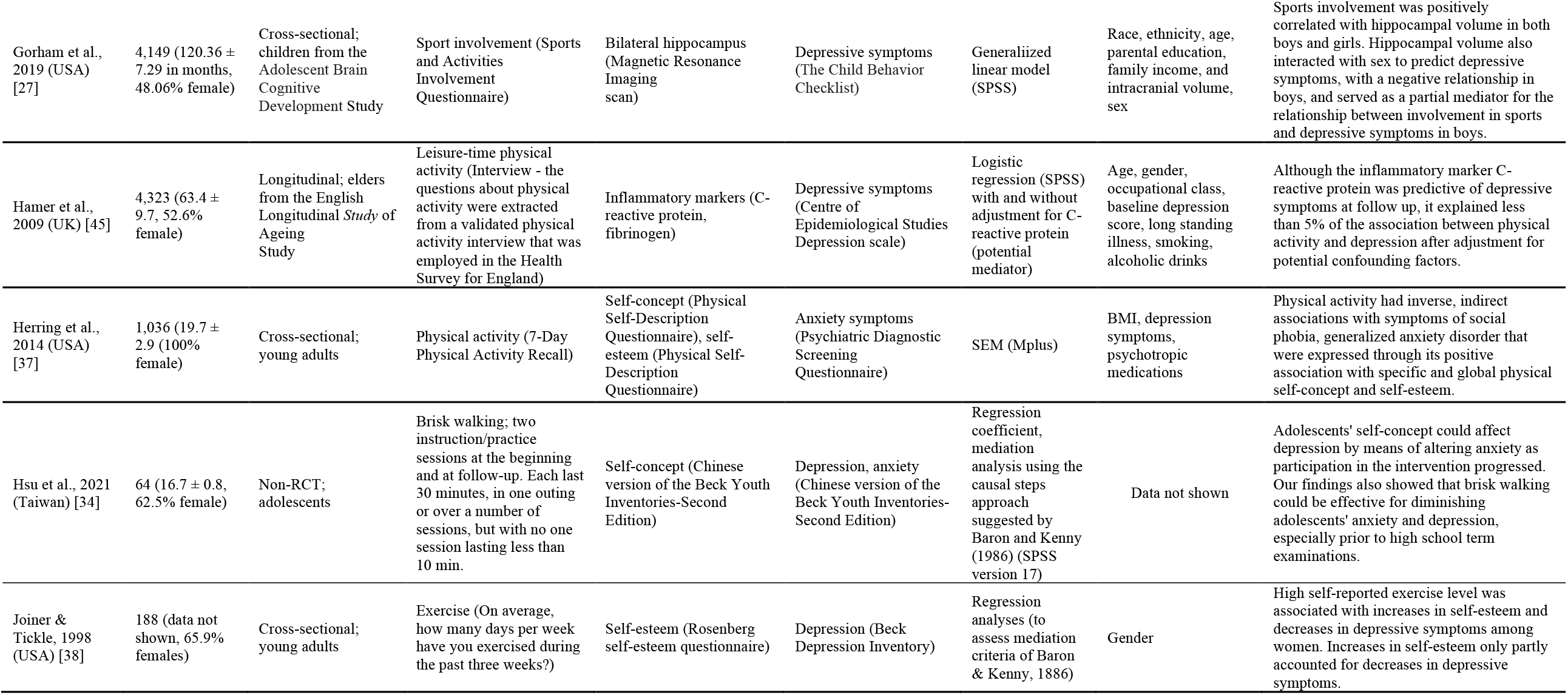
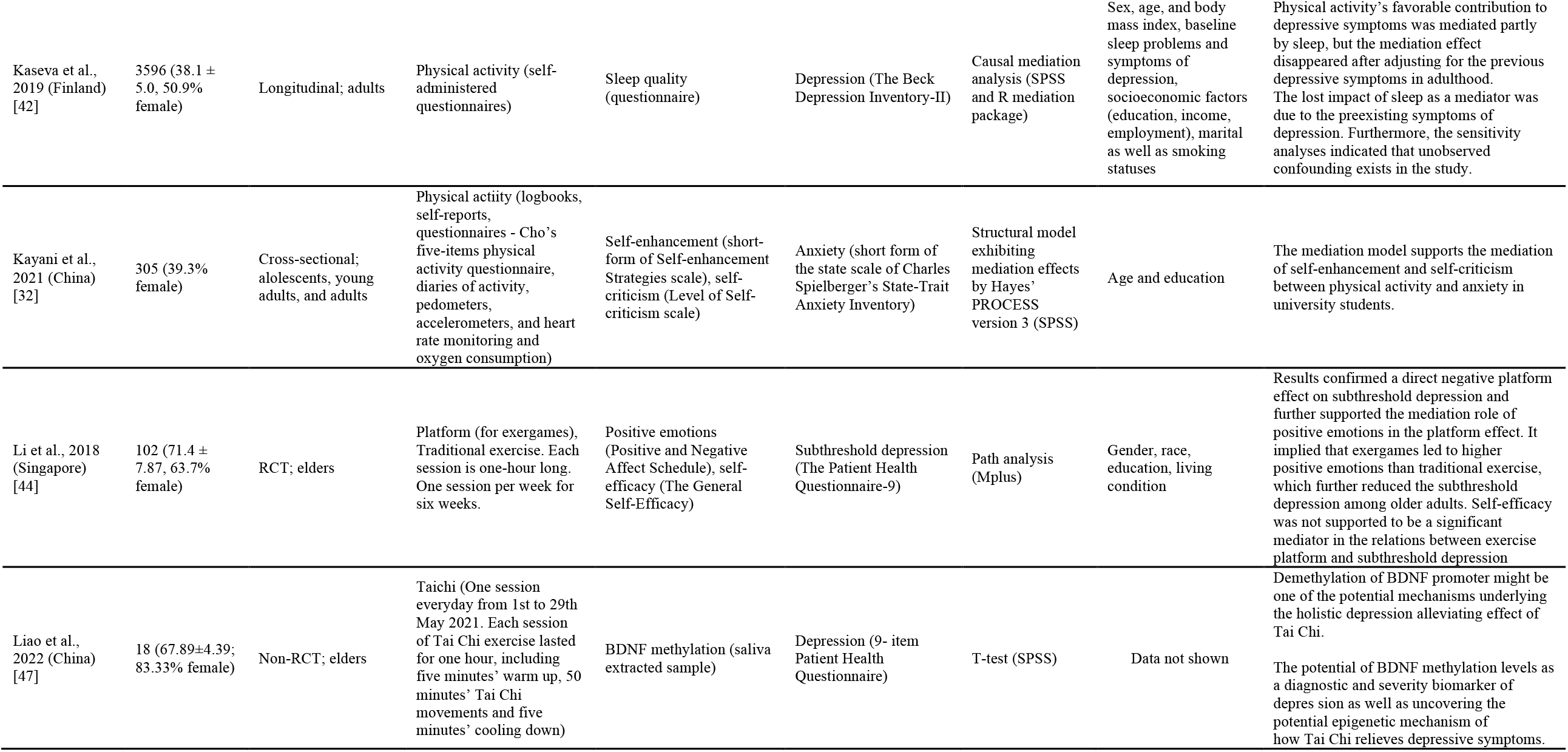
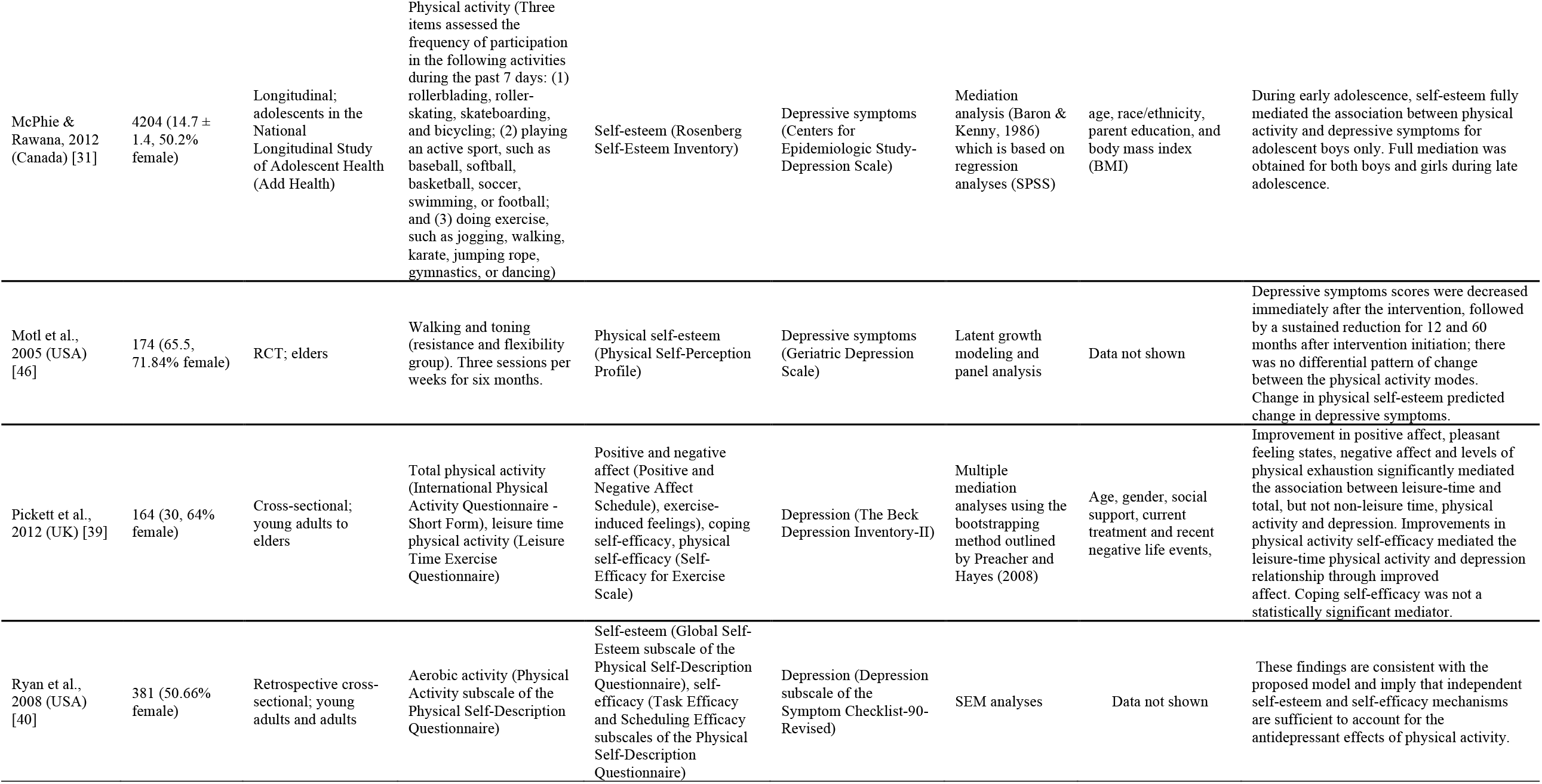
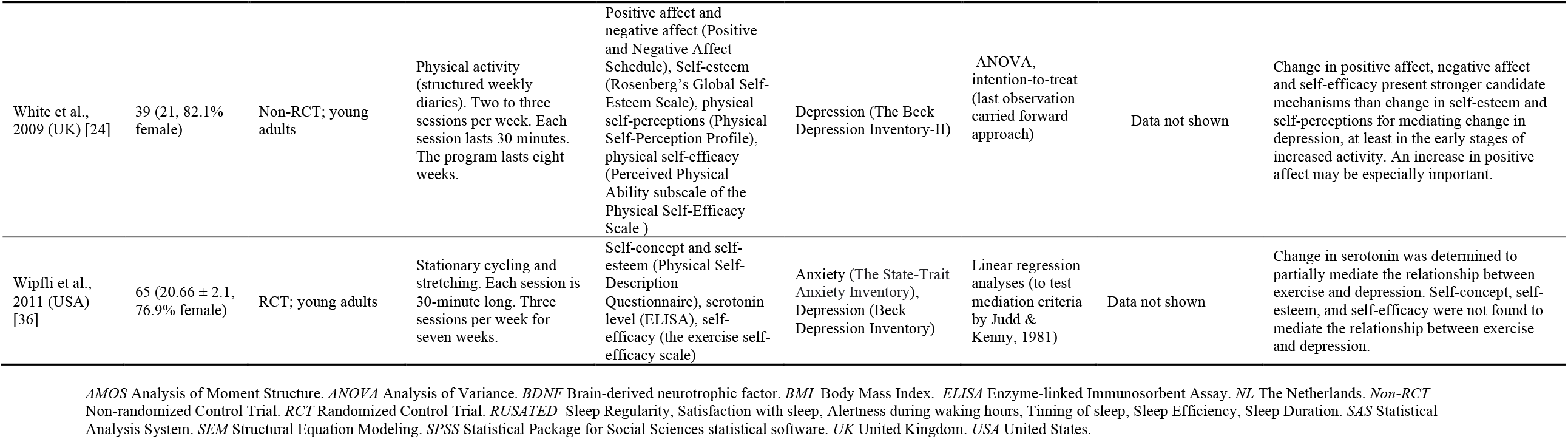
Summary of studies investigating the mediator of physical activity and psychiatric symptoms

| Authors, year (country) | n sample (mean age ± SD, % females) | Design; target population | Independent variable (instrument) | Mediating variable (instrument) | Dependent variable (instrument) | Statistical analysis; software | Confounders | Main findings |
| --- | --- | --- | --- | --- | --- | --- | --- | --- |
| Alghdir & Gabr, 2020 (Kingdom of Saudi Arabia) [43] | 80 (69.6 ± 5.38, 37.5% female) | Non-RCT; elders | Leisure-time physical activity. Each session is 45 to 60 minutes and consists of warm-up, active, and cool down phases. Three sessions per week for 12 weeks. | Adrenal hormones | Depressive mood (Profile of Mood States) | Multiple stepwise regressions, Pearson's correlation (SPSS) | Data not shown | The findings showed that 12 weeks of supervised exercise interventions are promising non-drug therapeutic strategies in improving depression among older adults. The potential performance in a psychological state occurs physiologically via optimizing the levels of the hormones of the hypothalamus-pituitary-adrenal axis. |
| Ansari et al., 2011 (UK) [33] | 3705 (77.9% female) | Cross-sectional; aolescents, young adults, and adults | Physical activity (survey) | Body image perception (Students were asked: "In your opinion are you...", with five response options ("Far too thin", "A little too thin", "Just right", "A little overweight", "Very overweight"). For the analysis, the five options were re-coded into three binary variables ("Underweight", "Just right", "Overweight").), self-rated health status ("How would you rate your health in general?") | Depression (Modified Beck Depression Inventory) | Regression, stratification analysis (SPSS) | Gender, perceived health, health awareness, muscle strengthening, academic performance | The positive effect of physical activity on depression could be down modulated by the negative impact of a 'distorted' body image on depression. While low physical activity level was not associated with depression, the protective effect of physical activity increased with physical activity intensity. The association between physical activity and depression may increase with physical activity intensity. |
| Babiss & Gangwisch, 2009 (USA) [25] | 14,594 (data not shown, 48.8% female) | Cross-sectional; participants in Wave 1 of the National Longitudinal Study of Adolescent Health (Add Health), adolescents | Sport participation (During the past week, how many times did you do exercise, such as jogging, walking, karate, jumping rope, gymnastics or dancing?) | Social support, self-esteem | Depression (18-item version of the Centers for Epidemiologic Study-Depression Scale) and suicidal ideation (subjects' yes or no responses to the question: "During the past 12 months, did you ever seriously think about committing suicide?") | Multivariate hierarchical logistic regression (SAS) | Sex, age, race/ethnicity, receipt of public assistance, and presence of physical limitations that could limit or preclude participation in sports and exercise | The results are consistent with self-esteem and social support acting as partial mediators of the relationships among sports participation and depression. As the adolescents' participation in sports increased, their average self-esteem was progressively higher, and those with higher self-esteem were less likely to suffer from depression. |
| Barham et al., 2021 (USA) [41] | 1576 (39.3 ± 13.7, 40% female) | Cross-sectional; adults | Physical activity (questionnaire, a single-item self-report measure) | Sleep health (the 6-question RUSATED Sleep Health survey V2.0) | Depression (Depression subscale of the 14-item Hospital Anxiety and Depression Scale) | Ordinary least squares regression, PROCESS macro (SPSS) | Sex, age, subjective social status, race, neuroticism, smoking status, chronotype, and self-reported cardiometabolic diagnoses or medication use | Sleep health was a significant predictor of depression symptoms and significantly mediated 19% of the association between physical activity scores and depression symptoms, even after controlling for sociodemographic, behavioral, and health-related covariates. |
| Booij et al., 2015 (NL) [29] | 715 (13.5 ± 0.5, 50.9% female) | Longitudinal; adolescents from the Tracking Adolescents' Individual Lives Survey study with three-year follow-up | Exercise (Participants could specify up to four different types of exercise that they regularly performed, and indicate for every exercise how many days and hours per week they spent on those activities) | Cortisol and heart rate responses to stress (Groningen Social Stress Test), C-reactive protein (immunonephelometric method) | Depressive symptoms (Affective Problems Scale of the Youth Self-Report, a somatic symptoms subscale and an affective symptoms subscale) | Linear regression, indirect method (Preacher & Hayes, 2008) | Gender, social economic status, BMI, total affective or somatic symptom scores, smoking, oral contraceptive use | Exercise was prospectively and inversely related to affective but not somatic symptoms. Heart rate during social stress partially mediated this relationship. No other mediating effects were found. |
| Chae et al., 2017 (South Korea) [35] | 848 (data not shown, 58.3% female) | Cross-sectional; adolescents | Physical activity (Physical Activity Questionnaire - Adolescent) | Self-esteem (Body-Esteem Scale For Adolescents and Adults) | Depression (Korean version of Children's Depression Inventory) | Path analysis, structural equation modeling, AMOS (SPSS) | Gender | Girls showed a significantly higher level of depressive symptoms than boys. Boys showed significantly higher levels of physical activity and body-esteem than girls. Body-esteem mediated the relation of physical activity with depression. |
| Conley et al., 2020 (USA) [28] | 4861 (9.8 ± 0.6, 47.5% female) | Cross-sectional; adults and children | Social physical activity (Adolescent Brain Cognitive Development Sports Activities Involvement Questionnaire) | Social connections (Adolescent Brain Cognitive Development Other Resilience Scale) | Depressive symptoms (The Child Behavior Checklist) | Causal mediation analysis (R mediation package) | Race, age in months, sex, income, site, and family | Number of close friends partially mediated (5.4%, p<0.01) the influence of social physical activity on depressive symptoms. These analyses were replicated in a higher sample of children from the Adolescent Brain Cognitive Development Study (n=7355). |
| Dishman et al., 2006 (USA) [30] | 1,250 (17.66 ± 0.61, 100% female) | Cross-sectional; adolescents | Physical activity (3-Day Physical Activity Recall) and sport participation (was calculated as the sum of two items adapted from the Youth Risk Behavior Surveillance Survey) | Self-concept (The Physical Self-Description Questionnaire) | Depression symptoms (The Center for Epidemiological Studies—Depression Scale) | SEM (AMOS) | Physical fitness, BMI | The results provide initial evidence suggesting that physical self-concept mediates the relations of physical activity and sport participation with self-esteem, which is inversely related to depression symptoms among girls in late adolescence. |
| Gorham & Barch, 2020 (USA) [26] | 3,973 (120.55 ± 7.32 in months, 48.2% female) | Cross-sectional; children from the Adolescent Brain Cognitive Development Study | Sport involvement (Sports and Activities Involvement Questionnaire) | White matter tract integrity (fractional anisotropy), Diffusion Magnetic Resonance Imaging metrics) | Depression (The Child Behavior Checklist) | Generalized linear model | Sex, race, ethnicity, age, parental education, and family income | Involvement in sports was associated with reduced depression in boys. The number of activities and sports that a child was involved in was negatively related to fractional anisotropy of the left fornix but was unrelated to fractional anisotropy of other tracts. fractional anisotropy of these white matter tracts was also unrelated to depressive symptoms. |
| Gorham et al., 2019 (USA) [27] | 4,149 (120.36 ± 7.29 in months, 48.06% female) | Cross-sectional; children from the Adolescent Brain Cognitive Development Study | Sport involvement (Sports and Activities Involvement Questionnaire) | Bilateral hippocampus (Magnetic Resonance Imaging scan) | Depressive symptoms (The Child Behavior Checklist) | Generalized linear model (SPSS) | Race, ethnicity, age, parental education, family income, and intracranial volume, sex | Sports involvement was positively correlated with hippocampal volume in both boys and girls. Hippocampal volume also interacted with sex to predict depressive symptoms, with a negative relationship in boys, and served as a partial mediator for the relationship between involvement in sports and depressive symptoms in boys. |
| Hamer et al., 2009 (UK) [45] | 4,323 (63.4 ± 9.7, 52.6% female) | Longitudinal; elders from the English Longitudinal Study of Ageing Study | Leisure-time physical activity (Interview - the questions about physical activity were extracted from a validated physical activity interview that was employed in the Health Survey for England) | Inflammatory markers (C-reactive protein, fibrinogen) | Depressive symptoms (Centre of Epidemiological Studies Depression scale) | Logistic regression (SPSS) with and without adjustment for C-reactive protein (potential mediator) | Age, gender, occupational class, baseline depression score, long standing illness, smoking, alcoholic drinks | Although the inflammatory marker C-reactive protein was predictive of depressive symptoms at follow up, it explained less than 5% of the association between physical activity and depression after adjustment for potential confounding factors. |
| Herring et al., 2014 (USA) [37] | 1,036 (19.7 ± 2.9 (100% female) | Cross-sectional; young adults | Physical activity (7-Day Physical Activity Recall) | Self-concept (Physical Self-Description Questionnaire), self-esteem (Physical Self-Description Questionnaire) | Anxiety symptoms (Psychiatric Diagnostic Screening Questionnaire) | SEM (Mplus) | BMI, depression symptoms, psychotropic medications | Physical activity had inverse, indirect associations with symptoms of social phobia, generalized anxiety disorder that were expressed through its positive association with specific and global physical self-concept and self-esteem. |
| Hsu et al., 2021 (Taiwan) [34] | 64 (16.7 ± 0.8, 62.5% female) | Non-RCT; adolescents | Brisk walking; two instruction/practice sessions at the beginning and at follow-up. Each last 30 minutes, in one outing or over a number of sessions, but with no one session lasting less than 10 min. | Self-concept (Chinese version of the Beck Youth Inventories-Second Edition) | Depression, anxiety (Chinese version of the Beck Youth Inventories-Second Edition) | Regression coefficient, mediation analysis using the causal steps approach suggested by Baron and Kenny (1986) (SPSS version 17) | Data not shown | Adolescents' self-concept could affect depression by means of altering anxiety as participation in the intervention progressed. Our findings also showed that brisk walking could be effective for diminishing adolescents' anxiety and depression, especially prior to high school term examinations. |
| Joiner & Tickle, 1998 (USA) [38] | 188 (data not shown, 65.9% females) | Cross-sectional; young adults | Exercise (On average, how many days per week have you exercised during the past three weeks?) | Self-esteem (Rosenberg self-esteem questionnaire) | Depression (Beck Depression Inventory) | Regression analyses (to assess mediation criteria of Baron & Kenny, 1986) | Gender | High self-reported exercise level was associated with increases in self-esteem and decreases in depressive symptoms among women. Increases in self-esteem only partly accounted for decreases in depressive symptoms. |
| Kaseva et al., 2019 (Finland) [42] | 3596 (38.1 ± 5.0, 50.9% female) | Longitudinal; adults | Physical activity (self-administered questionnaires) | Sleep quality (questionnaire) | Depression (The Beck Depression Inventory-II) | Causal mediation analysis (SPSS and R mediation package) | Sex, age, and body mass index, baseline sleep problems and symptoms of depression, socioeconomic factors (education, income, employment), marital as well as smoking statuses | Physical activity's favorable contribution to depressive symptoms was mediated partly by sleep, but the mediation effect disappeared after adjusting for the previous depressive symptoms in adulthood. The lost impact of sleep as a mediator was due to the preexisting symptoms of depression. Furthermore, the sensitivity analyses indicated that unobserved confounding exists in the study. |
| Kayani et al., 2021 (China) [32] | 305 (39.3% female) | Cross-sectional; adolescents, young adults, and adults | Physical activity (logbooks, self-reports, questionnaires - Cho's five-items physical activity questionnaire, diaries of activity, pedometers, accelerometers, and heart rate monitoring and oxygen consumption) | Self-enhancement (short-form of Self-enhancement Strategies scale), self-criticism (Level of Self-criticism scale) | Anxiety (short form of the state scale of Charles Spielberger's State-Trait Anxiety Inventory) | Structural model exhibiting mediation effects by Hayes' PROCESS version 3 (SPSS) | Age and education | The mediation model supports the mediation of self-enhancement and self-criticism between physical activity and anxiety in university students. |
| Li et al., 2018 (Singapore) [44] | 102 (71.4 ± 7.87, 63.7% female) | RCT; elders | Platform (for exergames), Traditional exercise. Each session is one-hour long. One session per week for six weeks. | Positive emotions (Positive and Negative Affect Schedule), self-efficacy (The General Self-Efficacy) | Subthreshold depression (The Patient Health Questionnaire-9) | Path analysis (Mplus) | Gender, race, education, living condition | Results confirmed a direct negative platform effect on subthreshold depression and further supported the mediation role of positive emotions in the platform effect. It implied that exergames led to higher positive emotions than traditional exercise, which further reduced the subthreshold depression among older adults. Self-efficacy was not supported to be a significant mediator in the relations between exercise platform and subthreshold depression |
| Liao et al., 2022 (China) [47] | 18 (67.89±4.39; 83.33% female) | Non-RCT; elders | Taichi (One session everyday from 1st to 29th May 2021. Each session of Tai Chi exercise lasted for one hour, including five minutes' warm up, 50 minutes' Tai Chi movements and five minutes' cooling down) | BDNF methylation (saliva extracted sample) | Depression (9- item Patient Health Questionnaire) | T-test (SPSS) | Data not shown | Demethylation of BDNF promoter might be one of the potential mechanisms underlying the holistic depression alleviating effect of Tai Chi.<br>The potential of BDNF methylation levels as a diagnostic and severity biomarker of depression as well as uncovering the potential epigenetic mechanism of how Tai Chi relieves depressive symptoms. |
| McPhie & Rawana, 2012 (Canada) [31] | 4204 (14.7 ± 1.4, 50.2% female) | Longitudinal; adolescents in the National Longitudinal Study of Adolescent Health (Add Health) | Physical activity (Three items assessed the frequency of participation in the following activities during the past 7 days: (1) rollerblading, roller-skating, skateboarding, and bicycling; (2) playing an active sport, such as baseball, softball, basketball, soccer, swimming, or football; and (3) doing exercise, such as jogging, walking, karate, jumping rope, gymnastics, or dancing) | Self-esteem (Rosenberg Self-Esteem Inventory) | Depressive symptoms (Centers for Epidemiologic Study-Depression Scale) | Mediation analysis (Baron & Kenny, 1986) which is based on regression analyses (SPSS) | age, race/ethnicity, parent education, and body mass index (BMI) | During early adolescence, self-esteem fully mediated the association between physical activity and depressive symptoms for adolescent boys only. Full mediation was obtained for both boys and girls during late adolescence. |
| Motl et al., 2005 (USA) [46] | 174 (65.5, 71.84% female) | RCT; elders | Walking and toning (resistance and flexibility group). Three sessions per weeks for six months. | Physical self-esteem (Physical Self-Perception Profile) | Depressive symptoms (Geriatric Depression Scale) | Latent growth modeling and panel analysis | Data not shown | Depressive symptoms scores were decreased immediately after the intervention, followed by a sustained reduction for 12 and 60 months after intervention initiation; there was no differential pattern of change between the physical activity modes. Change in physical self-esteem predicted change in depressive symptoms. |
| Pickett et al., 2012 (UK) [39] | 164 (30, 64% female) | Cross-sectional; young adults to elders | Total physical activity (International Physical Activity Questionnaire - Short Form), leisure time physical activity (Leisure Time Exercise Questionnaire) | Positive and negative affect (Positive and Negative Affect Schedule), exercise-induced feelings), coping self-efficacy, physical self-efficacy (Self-Efficacy for Exercise Scale) | Depression (The Beck Depression Inventory-II) | Multiple mediation analyses using the bootstrapping method outlined by Preacher and Hayes (2008) | Age, gender, social support, current treatment and recent negative life events, | Improvement in positive affect, pleasant feeling states, negative affect and levels of physical exhaustion significantly mediated the association between leisure-time and total, but not non-leisure time, physical activity and depression. Improvements in physical activity self-efficacy mediated the leisure-time physical activity and depression relationship through improved affect. Coping self-efficacy was not a statistically significant mediator. |
| Ryan et al., 2008 (USA) [40] | 381 (50.66% female) | Retrospective cross-sectional; young adults and adults | Aerobic activity (Physical Activity subscale of the Physical Self-Description Questionnaire) | Self-esteem (Global Self-Esteem subscale of the Physical Self-Description Questionnaire), self-efficacy (Task Efficacy and Scheduling Efficacy subscales of the Physical Self-Description Questionnaire) | Depression (Depression subscale of the Symptom Checklist-90-Revised) | SEM analyses | Data not shown | These findings are consistent with the proposed model and imply that independent self-esteem and self-efficacy mechanisms are sufficient to account for the antidepressant effects of physical activity. |
| White et al., 2009 (UK) [24] | 39 (21, 82.1% female) | Non-RCT; young adults | Physical activity (structured weekly diaries). Two to three sessions per week. Each session lasts 30 minutes. The program lasts eight weeks. | Positive affect and negative affect (Positive and Negative Affect Schedule), Self-esteem (Rosenberg's Global Self-Esteem Scale), physical self-perceptions (Physical Self-Perception Profile), physical self-efficacy (Perceived Physical Ability subscale of the Physical Self-Efficacy Scale ) | Depression (The Beck Depression Inventory-II) | ANOVA, intention-to-treat (last observation carried forward approach) | Data not shown | Change in positive affect, negative affect and self-efficacy present stronger candidate mechanisms than change in self-esteem and self-perceptions for mediating change in depression, at least in the early stages of increased activity. An increase in positive affect may be especially important. |
| Wipfli et al., 2011 (USA) [36] | 65 (20.66 ± 2.1, 76.9% female) | RCT; young adults | Stationary cycling and stretching. Each session is 30-minute long. Three sessions per week for seven weeks. | Self-concept and self-esteem (Physical Self-Description Questionnaire), serotonin level (ELISA), self-efficacy (the exercise self-efficacy scale) | Anxiety (The State-Trait Anxiety Inventory), Depression (Beck Depression Inventory) | Linear regression analyses (to test mediation criteria by Judd & Kenny, 1981) | Data not shown | Change in serotonin was determined to partially mediate the relationship between exercise and depression. Self-concept, self-esteem, and self-efficacy were not found to mediate the relationship between exercise and depression. |

*AMOS* Analysis of Moment Structure. *ANOVA* Analysis of Variance. *BDNF* Brain-derived neurotrophic factor. *BMI* Body Mass Index. *ELISA* Enzyme-linked Immunosorbent Assay. *NL* The Netherlands. *Non-RCT* Non-randomized Control Trial. *RCT* Randomized Control Trial. *RUSATED* Sleep Regularity, Satisfaction with sleep, Alertness during waking hours, Timing of sleep, Sleep Efficiency, Sleep Duration. *SAS* Statistical Analysis System. *SEM* Structural Equation Modeling. *SPSS* Statistical Package for Social Sciences statistical software. *UK* United Kingdom. *USA* United States.

#### 3.2.1 Characteristics of included studies

The included studies have sample sizes ranging from 39 [24] to 14,594 [25]. Three studies included children [26–28], eight studies included adolescents [24,25,28–35], nine studies included young adults [24,32,33,36–40], six studies included adults [32,33,39–42], and six studies included elderly people [39,43–47].

#### 3.2.2 Exposure characteristics

Out of 24 studies, four studies used sports participation [25–27,30], five studies used exercise [29,36,38,44,47], and 16 studies used physical activity as the predictor [24,28,30–35,37,39–43,45,46]. Three intervention studies included an active control group [36,44,46]. Within these studies, the experimental conditions included exergames [44], resistance and flexibility exercise [46], and cycling [36]. Three intervention studies did not have a control group; all participants had to participate in the same training program [34,43,47]. Session duration ranged from 30 minutes [36] to 60 minutes [43,44,47]. The frequency of the session per week ranged from one [44] to seven [47] times per week. Lastly, the overall duration of the interventions vary between four weeks [47] to 24 weeks [46].

#### 3.2.3 Mechanism characteristics

Within the 17 studies that explored the psychosocial mechanisms linking physical activity with psychiatric symptoms, two studies used social support/connection [25,28], one study used body image [33], one study used self-criticism and self-enhancement [32], eight studies used self-esteem [24,25,31,35–37,40,46], four studies used self-concept [30,34,36,37], five studies used self-efficacy [24,36,39,41,44], one study used self-perception [24] and three studies used mood [24,39,44]. as the potential mechanisms of interest. Among the seven studies that explored the neurobiological mechanisms, one study used hippocampal volume [28], one study used white matter microstructure [26], one study used stress-induced biomarkers [29], one study used neurotransmitters [36], one study used hormones [43], two studies used inflammatory biomarkers [29,45], one study used epigenetics [47]. The two studies that explored behavioral mechanisms focused on sleep [41,42].

#### 3.2.4 Outcomes characteristics

There were 22 studies that used depression as the outcome variable [24–31,33–36,38–46]. One study used the Profile of Mood Sates [43], three studies used the Center for Epidemiological Studies—Depression Scale [30,31,45], one study used the 18-item version of the Center for Epidemiologic Study-Depression Scale [25], one study used the Affective Problems Scale of the Youth Self-Report [29], one study used the Korean version of Children’s Depression Inventory [35], three studies used the Child Behavior Checklist [26–28], two studies used the Beck Depression Inventory [36,38], one study used The Patient Health Questionnaire-9 LI, one study used the Geriatric Depression Scale [46], three studies used the Beck Depression Inventory-II [24,39,42], one used the Chinese version of the Beck Youth Inventories-Second Edition [34], one study used the Depression subscale of the Hospital Anxiety and Depression Scale [41], one study used the Modified Beck Depression Inventory [33], one study used the Depression subscale of the Symptom Checklist-90-Revised [40], one study used the 9-item Patient Health Questionnaire [47]. One study also assessed the affective and somatic symptoms of depression [29].

Four studies used anxiety symptoms as the outcome variable [32,34,36,37]. In particular, one study used the State-Trait Anxiety Inventory [36], one study used the short form of the State scale from the Charles Spielberger’s State - Trait Anxiety Inventory [32], one used the Chinese version of the Beck Youth Inventories - Second Edition [34], and the other one the Psychiatric Diagnostic Screening Questionnaire [37].

### 3.3 Synthesis of findings

The qualitative synthesis of our findings was summarized in **Table 2**. Overall, there was consistent evidence for three mechanisms through which physical activity seems to have a link with psychiatric symptoms: self-esteem (eight out of 10 studies – 80%), self-concept (three out of four studies – 75%), and self-efficacy (three out of five studies – 60%). There were insufficient studies (less than four studies) to determine the role of social support, body image, self-criticism, self-enhancement, self-perception, mood, hippocampal volume, white matter microstructure, stress-induced markers, neurotransmitters, hormones, inflammatory markers, epigenetics, or sleep in the relationship between physical activity and psychiatric symptoms across the lifespan.

**Table 2.**
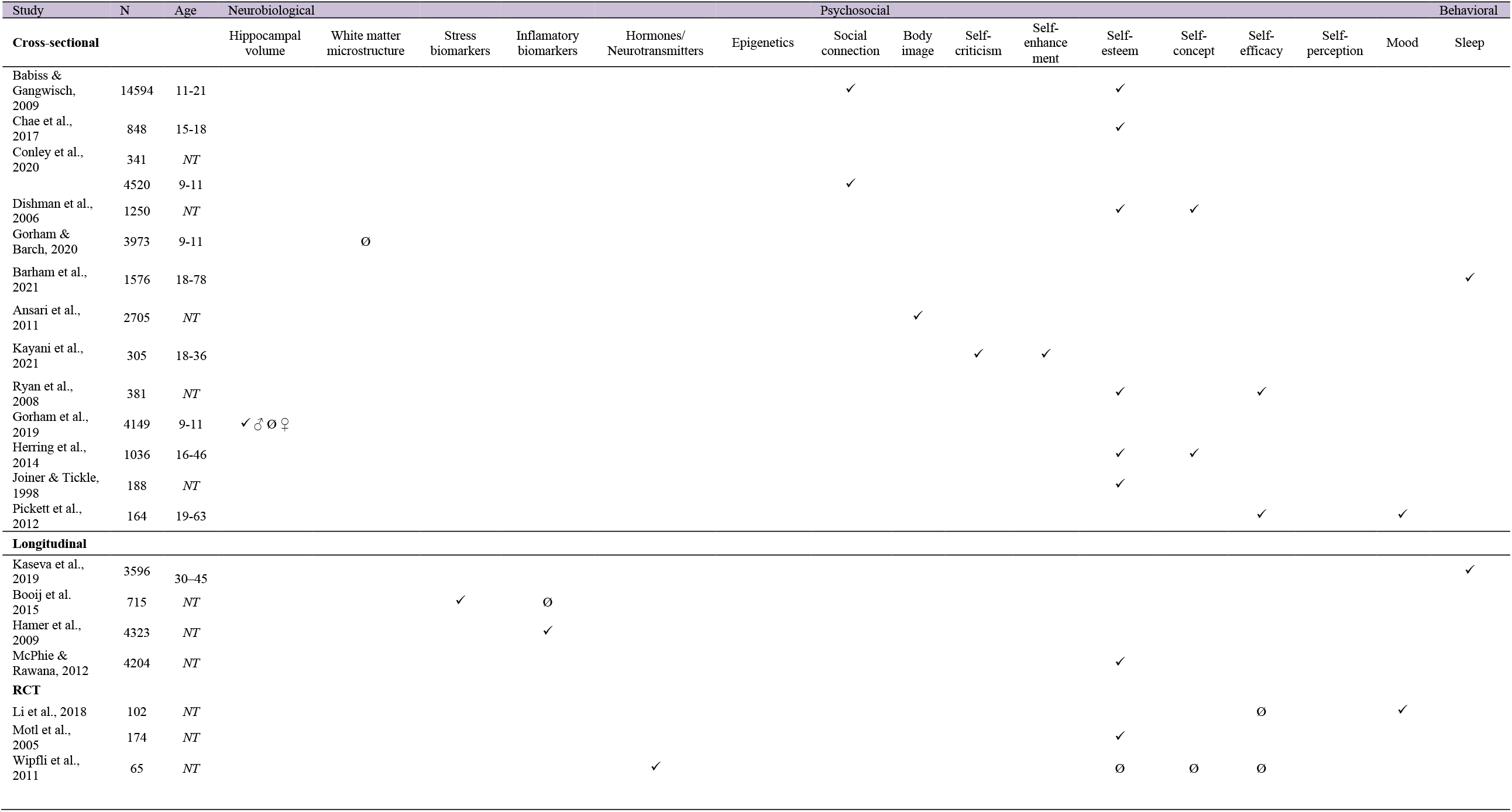
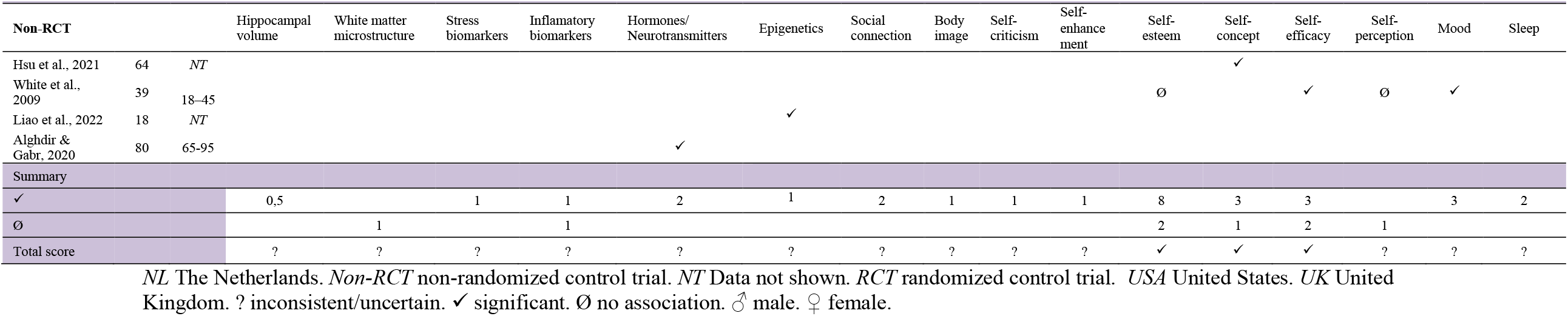
Qualitative synthesis

### 3.4 Risk of bias assessment

The details of the risk of bias assessment are included **Tables S2-S9**. The criteria to assess the risk of bias and the percentage of studies that met the criteria item-by-item is presented in **Tables S2-S5**. The risk of bias assessment study-by-study is presented in **Tables S6-S9**. First, all three RCTs showed high risk of bias [36,44,46]. Second, a high risk of bias was detected in two of the four non-RCTs [24,47]. Third, three out of four longitudinal studies showed low risk of bias [31,42,45]. Lastly, of the 13 cross-sectional studies, 11 showed high risk of bias [26–28,30,33,35,37–41], and two studies showed low risk of bias [25,32].

## 4. Discussion

The current systematic review aimed to synthesize research evidence on the mechanisms through which physical activity might reduce psychiatric symptoms across the lifespan. In brief, most of the studies focused on psychosocial mechanisms and consistently showed that self-esteem, self-concept, and self-efficacy are pathways through which physical activity reduces internalizing symptoms (i.e., depressive symptoms) in the healthy general population, and mainly in young people. We found that only a limited number of studies have explored the role of neurobiological (e.g., gray matter volume in the hippocampus) and behavioral (e.g., sleep) mechanisms in youth, making it difficult to obtain the overall picture. Therefore, future studies are encouraged to focus on: (i) exploring the neurobiological and behavioral mechanisms linking physical activity with psychiatric symptoms, and (ii) building a comprehensive and integrative model, that includes all potential mechanisms.

While a variety of psychosocial mechanisms were identified in this systematic review, only self-esteem, self-concept, and self-efficacy were consistently associated with physical activity and psychiatric symptoms. In brief, self-concept is understood as the beliefs and knowledge that we have about ourselves while self-esteem includes the evaluation and affect that we have regarding our self-concept [48]. On the other hand, self-efficacy is the belief in oneself to perform a certain action and achieving a particular result. Overall, our findings are in line with those from Kandola et al. (2019) and Lubans et al. (2016) who proposed that self-esteem may be a potential psychosocial mechanism through which physical activity can reduce depression [14,17]. More specifically, participation in physical activity may enhance one’s self-esteem through improving physical self-concept and body image perceptions, which in turn, might have a positive effect on psychiatric symptoms. While self-esteem has been often associated with depression, it could also play a role in symptoms of externalizing disorders such as risky behavior, aggression, and violence [48]. Therefore, future research might also investigate the role of self-esteem in reducing externalizing symptoms. Lastly, it was hypothesized that by doing physical activity, we will gain more confidence in our physical ability (i.e., perceived competence); hence, promote our self-efficacy. This improvement in self-efficacy will then generalize to other aspects and act against depressive symptoms. In addition to these mechanisms, other potential psychosocial mechanisms such as social support, mood and emotions may also mediate the relationship between physical activity and psychiatric symptoms [14,17]. For example, having a positive experience while doing physical activity may have a good influence on various psychosocial scales such as self-esteem, which in turn, affects mental health outcomes [49–51]. Indeed, some studies found evidence for other psychosocial outcomes (i.e., social connections, body image, self-criticism, self-enhancement, self-perception, and mood) [24,25,28,30,32,33,36,39,44]. However, there were not enough studies to draw strong conclusions. Accordingly, future research should focus on investigating these potential mechanisms and employing self-esteem, self-concept, and self-efficacy to develop effective physical activity interventions for mental health.

The role of the neurobiological mechanisms in the relationship between physical activity and psychiatric symptoms is unclear due to the inconsistencies and heterogeneity found between the included studies. For instance, some studies used magnetic resonance imaging (MRI) data as indicators of the neurobiological mechanisms while others included blood circulating neurobiological biomarkers [26,43]. Studies that used MRI data provided evidence for potential neurobiological mechanisms; however, the effects were inconsistent. Two studies that used the data from the Adolescent Brain and Cognitive Development (ABCD) study, explored the role of brain structure in the relationship between physical activity and psychiatric symptoms in children [26,27]. Overall, they observed that hippocampal volume partly mediated the association of physical activity with depressive symptoms, but only in boys [27]. However, white matter microstructure in the fornix and parahippocampal cingulum did not mediate the relationship between physical activity and psychiatric symptoms during childhood [26]. It is notable that the sample size of the ABCD study was quite large and hence could result in more robust results than expected.

In healthy young individuals, neurobiological measurements in the form of blood circulating biomarkers could provide a more dynamic indication of the role of neurobiological mechanisms in the relationship between physical activity and psychiatric symptoms across the lifespan. For example, stress and inflammatory blood biomarkers, which have been previously linked to psychiatric symptoms, can be reduced by physical activity interventions [29,45]. Psychiatric disorders are often associated with elevated level of stress and inflammatory biomarkers while physical activity has an anti-inflammatory effect. It is possible, therefore, that stress and inflammatory biomarkers could be a potential mechanism through which physical activity reduces psychiatric symptoms [17]. Similarly, adrenal hormones and neurotransmitters, such as the serotonin, are other potential neurobiological mechanisms that might be stimulated by physical activity, and in turn reduce depressive symptoms [36,43]. Lastly, physical activity may also affect psychiatric symptoms through epigenetic mechanisms [52]. For instance, Liao et al. (2022) explored the possibility that BDNF methylation could mediate the effect of physical activity on depressive symptoms in older people and found some preliminary evidence for this hypothesis [47]. Overall, although there are promising and highly accepted neurobiological mechanisms in the field, we cannot establish any conclusion in this systematic review about their potential role in the relationship between physical activity and psychiatric symptoms across the lifespan.

In 2016, Lubans et al. [14] described a range of potential behavioral mechanisms (e.g., sleep, coping, and regulation skills) that may explain the effects of physical activity on mental health during childhood. However, they only identified two studies that had tested behavioral mechanisms. Similarly, we found a lack of relevant studies examining behavioral mechanisms because most studies understood behavioral mechanisms as independent predictors of psychiatric symptoms instead of potential paths. In this study, we found and included two studies that identified sleep as a potential path through which physical activity might reduce psychiatric symptoms in adults ([41,42]. Previous studies showed that physical activity has a beneficial effect on sleep, where it can improve sleep quality and duration; additionally, sleep is essential for recovery after doing physical activity [42,53]. Furthermore, the relationship between sleep and psychiatric disorders, particularly depression, could be bidirectional [54]. This means that sleep could be a consequence as well as a predictor of some psychiatric symptoms. Nevertheless, there were insufficient number of studies to draw firm conclusions.

### 4.1 Literature gaps and future research

- While we aimed to investigate the mechanisms through which physical activity may influence externalizing and internalizing symptoms, we failed to identify any studies examining the effect of physical activity on externalising symptoms.
- The variety of measurements available for assessing depressive and anxiety symptoms may pose as an obstacle for research and clinical application. Researchers should consider using similar measures when assessing symptoms of the same disorder.
- Only three mechanisms (i.e., self-esteem, self-concept, self-efficacy) were sufficiently studied. Therefore, further research is needed to explore whether other psychosocial (e.g., social support), neurobiological and behavioral paths exist.
- Since most studies focused on children and adolescents, there is a need for additional studies examining potential mechanisms in adult and elderly populations.
- Integrated studies that examine the combined and independent contributions of neurobiological, psychosocial, and behavioral mechanisms are needed to obtain the overall picture.

### 4.2 Limitations and strengths

This review has a few limitations. First, conference abstracts and other types of grey literature were not included. Additionally, we only included articles published in English and did not perform a formal search for behavioral mechanisms due to feasibility reasons. Instead, we included articles that we found from screening the reference list of articles that met the inclusion criteria and contacted experts in the field. Finally, due to the heterogeneity of studies, we could not perform a meta-analysis. On the other hand, this review has a few strengths. We included participants of all ages and all types of studies. This review is an a priori study which was registered in the PROSPERO database [19]. We also followed the PRISMA guidelines for systematic review and included articles across five electronic databases **Table S1**.

## 5. Conclusion

Findings from our systematic review suggest that self-esteem, self-concept, and self-efficacy are potential paths through which physical activity might reduce psychiatric symptoms (specifically depressive and anxiety symptoms) across the lifespan. Therefore, future interventional studies should consider incorporating these mechanisms to develop more effective interventions. There were insufficient studies to establish the role of other psychosocial, neurobiological and behavioral mechanisms linking physical activity with psychiatric symptoms across the lifespan.

## Data Availability

All data produced in the present work are contained in the manuscript

## Statements and Declarations

### Availability of data and material

Not applicable

### Ethics approval

Not applicable

### Consent for publication

Not applicable

### Code availability

Not applicable

### Contributors

MR-A, PTNH, WMB, TPBH, TT designed and drafted the systematic review. WMB performed the search strategy. MR-A, PTNH, TPBH, TT, AH, DRL and MV revised and approved the final version of the manuscript. MR-A will be the guarantor of the review.

