## Supplementary material for "Mechanisms linking physical activity with psychiatric symptoms across lifespan: A systematic review"

María Rodriguez-Ayllon

Department of Epidemiology

Erasmus MC University Medical Center

Rotterdam, the Netherlands

**Table S1.** Excluded articles (n=149)

| Reasons | Number of articles | References |
|---|---|---|
| Wrong population | 8 | Bonura, K.B. & Tenenbaum, G. (2014); Chung, S.C, et al. (2012); Duda. J.L., et al. (2014); Gohr Månsson, A., et al. (2019); Miyata, H., et al. (2020); Narasingharao, K., et al. (2017); Ramanathan, M., et al. (2017); Furtado, G.E., et al. (2021) |
| Wrong intervention | 13 | Carter, J. S., et al. (2017); Haden, S.C., et al. (2014); Hilyer, J.C., et al. (1982); Luna, P., et al. (2019); Luna, P., et al. (2020); Mehta, P. & Sharma, M. (2011); Novaes, M.M., et al. (2020); Opdenacker, J., et al. (2008); Polku, H., et al. (2015); Riley, K.E., et al. (2017); Shin, K.R., et al. (2009); Van Hoecke, A.S., et al. (2014), Pereira et al., (2013) |
| Wrong outcome | 13 | Awick, E.A., et al. (2017); Benvenutti, M.J., et al. (2017); Bunketorp Käll, L., et al. (2015); Evans, M., et al. (2017); Fernandes, H.M. (2018); Jin, P., (1989); Malathi, A. & Damodaran, A. (1999); Napoli, N., et al. (2014); Xie, X., et al. (2021); Baykose, N., et al. (2021); Lyu, J., et al. (2021) ; Ng-Knight, T., et al. (2021) ; Shang, Y., et al. (2021) |
| Wrong study design | 3 | Archer, T. & Kostrzewa, R.N. (2012); Chan, S.H.W. & Tsang, H.W.H. (2019); Foreyt, J.P., et al. (1995) |
| No/wrong mediator | 108 | Paoluccia, E.M., et al. (2018); Chen, YC., et al. (2019); Aguinaga, S., et al. (2018); Alfermann, D. & Stoll, O. (2000); Antunes, H.K.M, et al. (2005); Aras, D. & Ewert, A.W. (2016); Asci, F.H. (2003); Aşçi, F.H. (2009); Ashdown-Franks, G., et al. (2017); Bartels, L., et al. (2019); Beets, M.W. & Mitchell, E. (2010); Benavides, S. & Caballero, J. (2009); Binsinger, C., et al. (2006); Bonhauser, M., et al. (2005); Brière, F.N., et al. (2020); Brière, F.N., et al. (2018); Broman-Fulks, J.J., et al. (2004); Camacho, T.C., et al. (1991); Cassilhas, R.C., et al. (2010); Chen, H.M., et al. (2016); Crews, D.J., et al. (2004); Danielly, Y. & Silverthorne, C. (2017); Daukantaite, D., et al. (2018); Draper, N., et al. (2012); Fakhari, M. (2017); Farzane, A. & Koushkie Jahromi, M. (2021); Feiss, R. & Pangelinan, M.M. (2021); Fidelix, Y., et al. (2019); Gammage, K.L., et al. (2016); Guimaraes, T.T., et al. (2015); Gujral, S., et al. (2014); Guo, F., et al. (2020); Haas, P., et al. (2017); Hallgren, M., et al. (2020); Hawker, C.L. (2012); Heesch, K.C., et al. (2015); Hong, X., et al. (2009); Hoying, J. & Melnyk, B.M. (2015); Huang, T.T., et al. (2015); Ishii, K., et al. (2016); Jambor, E.A., et al. (1994); Jarry, J.L., et al. (2017); Katula, J.A., et al. (1999); Khorvash, M., et al. (2012); Kim, K.B., et al. (2004); Kim, Y.M. & Cho, S.I. (2021); Kim, Y.S., et al. (2019); Kleppang, A.L., et al. (2018); Kliziene, I., et al. (2018); Knippenberg, I.A.H., et al. (2021); Langguth, A.N., et al. (2016); Lee, M.S., et al. (2004); Legrand, F.D. (2014); Lerche, S., et al. (2018); Lobstein, D.D. & Rasmussen, C.L. (1991); Lobstein, D.D., et al. (1989); MacMahon, J.R. & Gross, R.T. (1988); Marquez, D.X., et al. (2002); Marselle, M.R., et al. (2014); McMahon, K., et al. (2021); McPhie, M.L. & Rawana, J.S. (2015); Meier, N.F. & Welch, A.S. (2016); Mendonça, R.M.S.C., et al. (2015); Moeijes, J., et al. (2019); Moses, J., et al. (1989); Mulcahy, A., et al. (2020); Nabkasorn, C., et al. (2006); O'Toole, S., et al. (2018); Palenzuela, D.L., et al. (2018); Palmer, L.K. (1995); Palmer, Y. (2014); Papp, M.E., et al. (2019); Partonen, T., et al. (1998); Petty, K.H., et al. (2009); Pisarska, A., et al. (2018); Roh, H.T., et al. (2018); Romero-Perez, E.M., et al. (2020); Sampasa-Kanyinga, H., et al. (2020); Shepherd, D., et al. (2012); Smith, J.A., et al. (2011); Streeter, C.C., et al. (2010); Taspinar, B., et al. (2014); Tejvani, R., et al. (2016); Thordardottir, K., et al. (2016); Tsutsumi, T., et al. (1997); van Gool, C.H., et al. |

| | | |
|---|---|---|
| | | (2007); van Heuvelen, M.J.G., et al. (2005); Vernon, J.B., et al. (2018); Viana, R.B., et al. (2019); Wang, S.K., et al. (2018); Wheatley, C., et al. (2020); White, K., et al. (2009); Williams, C.F., et al. (2019); Wise, L.A., et al. (2006); Xu, W., et al. (2017); Yeh, S.H., et al. (2015); Yigiter, K. & Hardee, J.T. (2017); Yoshihara, K., et al. (2014); Fink, A., et al. (2021) ; Forseth, B., et al. (2021) ; Ge, L.K., et al. (2021) ; Hofman, A., et al. (2021) ; Kang, H. & Jang, S. (2021) ; Ray, U.S., et al. (2001); Sheppard, A.C. & Mahoney, J.L. (2012); Wadhen, V. & Cartwright, T. (2021); Lozano Montes, L., et al. (2021); Forseth, B., et al. (2022) |
| No fulltext | 4 | Fasting, K. & Gronningsaeter, H. (1986); Li, X.Z. (2005); Liu, B.Q. & Qiu, Y. (2007); Ojha, H. & Yadav, N.P. (2016) |

**Table S2.** Criteria for methodological risk of bias assessment of included randomized control trial articles and percentage of studies meeting these criteria.

| Criteria items | Percentage of studies meeting criterion (%) |
|---|---|
| 1. Was true randomization used for assignment of participants to treatment groups? | 33.3 |
| 2. Was allocation to treatment groups concealed? | 0 |
| 3. Were treatment groups similar at the baseline? | 66.7 |
| 4. Were participants blind to treatment assignment? | 0 |
| 5. Were those delivering treatment blind to treatment assignment? | 0 |
| 6. Were outcomes assessors blind to treatment assignment? | 0 |
| 7. Were treatment groups treated identically other than the intervention of interest? | 100 |
| 8. Was follow up complete and if not, were differences between groups in terms of their follow up adequately described and analyzed? | 100 |
| 9. Was true randomization used for assignment of participants to treatment groups? | 25 |
| 10. Were outcomes measured in the same way for treatment groups? | 100 |
| 11. Were outcomes measured in a reliable way? | 100 |
| 12. Was appropriate statistical analysis used? | 100 |
| 13.Was the trial design appropriate, and any deviations from the standard RCT design (individual randomization, parallel groups) accounted for in the conduct and analysis of the trial? | 100 |

Adapted from the Joanna Briggs Institute Critical Appraisal Tool for Systematic Reviews;
-: Not applicable criterion

**Table S3.** Criteria for methodological risk of bias assessment of included non-randomized control trial articles and percentage of studies meeting these criteria.

| Criteria items | Percentage of studies meeting criterion (%) |
|---|---|
| 1. Is it clear in the study what is the 'cause' and what is the 'effect' (i.e. there is no confusion about which variable comes first)? | 100 |
| 2. Were the participants included in any comparisons similar? | 100 |
| 3. Were the participants included in any comparisons receiving similar treatment/care, other than the exposure or intervention of interest? | 100 |
| 4. Was there a control group? | 25 |
| 5. Were there multiple measurements of the outcome both pre and post the intervention/exposure? | 100 |
| 6. Was follow up complete and if not, were differences between groups in terms of their follow up adequately described and analyzed? | 75 |
| 7. Were the outcomes of participants included in any comparisons measured in the same way? | 75 |
| 8. Were outcomes measured in a reliable way? | 100 |
| 9. Was appropriate statistical analysis used? | 50 |

Adapted from the Joanna Briggs Institute Critical Appraisal Tool for Systematic Reviews; - : Not applicable criterion

**Table S4.** Criteria for methodological risk of bias assessment of included longitudinal articles and percentage of studies meeting these criteria.

| Criteria items | Percentage of studies meeting criterion (%) |
|---|---|
| 1. Were the two groups similar and recruited from the same population? | 100 |
| 2. Were the exposures measured similarly to assign people to both exposed and unexposed groups? | 100 |
| 3. Was the exposure measured in a valid and reliable way? | 20 |
| 4. Were confounding factors identified? | 100 |
| 5. Were strategies to deal with confounding factors stated? | 100 |
| 6. Were the groups/participants free of the outcome at the start of the study (or at the moment of exposure)? | - |
| 7. Were the outcomes measured in a valid and reliable way? | 100 |
| 8. Was the follow up time reported and sufficient to be long enough for outcomes to occur? | 100 |
| 9. Was follow up complete, and if not, were the reasons to loss to follow up described and explored? | 100 |
| 10. Were strategies to address incomplete follow up utilized? | 100 |
| 11. Was appropriate statistical analysis used? | 100 |

Adapted from the Joanna Briggs Institute Critical Appraisal Tool for Systematic Reviews; - : Not applicable criterion

**Table S5.** Criteria for methodological risk of bias assessment of included cross-sectional articles and percentage of studies meeting these criteria.

| Criteria items | Percentage of studies meeting criterion (%) |
|---|---|
| 1. Were the criteria for inclusion in the sample clearly defined? | 38.5 |
| 2. Were the study subjects and the setting described in detail? | 46.2 |
| 3. Was the exposure measured in a valid and reliable way? | 76.9 |
| 4. Were objective, standard criteria used for measurement of the condition? | - |
| 5. Were confounding factors identified? | 53.8 |
| 6. Were strategies to deal with confounding factors stated? | 100 |
| 7. Were the outcomes measured in a valid and reliable way? | 100 |
| 8. Was appropriate statistical analysis used? | 100 |

Adapted from the Joanna Briggs Institute Critical Appraisal Tool for Systematic Reviews; - : Not applicable criterion

**Table S6.** Risk assessment of included randomized control trial articles.

| Study | Item 1 | Item 2 | Item 3 | Item 4 | Item 5 | Item 6 | Item 7 | Item 8 | Item 9 | Item 10 | Item 11 | Item 12 | Item 13 | Quality score % | Risk category |
|---|---|---|---|---|---|---|---|---|---|---|---|---|---|---|---|
| Li et al. 2018 | × | × | ✓ | × | × | × | ✓ | - | ✓ | ✓ | ✓ | ✓ | ✓ | 58.3 | High risk |
| Motl et al. 2005 | × | × | × | × | × | × | ✓ | ✓ | ✓ | ✓ | ✓ | ✓ | ✓ | 53.8 | High risk |
| Wipfli et al. 2011 | ✓ | × | ✓ | × | × | × | ✓ | - | ✓ | ✓ | ✓ | ✓ | ✓ | 66.7 | High risk |
| Criterion score % | 33,3 | 0 | 66,7 | 0 | 0 | 0 | 100 | 100 | 100 | 100 | 100 | 100 | 100 | | |

Note that the total risk of bias score was calculated by dividing the number of criteria met in one study by the total number of criteria (i.e., 10). Note that the criterion score is calculated by dividing the number of studies meeting one criterion by the total number of studies (i.e., 4). ✓: meet the methodological quality criterion; ×: not meet the methodological quality criterion; - Not applicable criterion.

**Table S7.** Risk assessment of included non-randomized control trial articles.

| Study | Item 1 | Item 2 | Item 3 | Item 4 | Item 5 | Item 6 | Item 7 | Item 8 | Item 9 | Quality score % | Risk category |
|---|---|---|---|---|---|---|---|---|---|---|---|
| Alghdir & Gabr 2020 | ✓ | ✓ | ✓ | ✓ | ✓ | ✓ | ✓ | ✓ | ✓ | 100 | Low risk |
| Hsu et al. 2021 | ✓ | - | - | × | ✓ | ✓ | ✓ | ✓ | ✓ | 85.7 | Low risk |
| White et al. | ✓ | - | - | × | ✓ | × | × | ✓ | × | 42.6 | High risk |
| Liao et al. | ✓ | - | - | × | ✓ | ✓ | ✓ | ✓ | × | 71.4 | High risk |
| Criterion score % | 100 | 100 | 100 | 25 | 100 | 75 | 75 | 100 | 50 | | |

Note that the total risk of bias score was calculated by dividing the number of criteria met in one study by the total number of criteria (i.e., 10). Note that the criterion score is calculated by dividing the number of studies meeting one criterion by the total number of studies (i.e., 4). ✓: meet the methodological quality criterion; ×: not meet the methodological quality criterion; - Not applicable criterion.

**Table S8.** Risk assessment of included longitudinal articles.

| Study | Item 1 | Item 2 | Item 3 | Item 4 | Item 5 | Item 6 | Item 7 | Item 8 | Item 9 | Item 10 | Item 11 | Quality score % | Risk category |
|---|---|---|---|---|---|---|---|---|---|---|---|---|---|
| Hamer et al. 2009 | ✓ | ✓ | ✓ | ✓ | ✓ | - | ✓ | ✓ | × | ✓ | ✓ | 90 | Low risk |
| McPhie & Rawana 2012 | ✓ | ✓ | × | ✓ | ✓ | - | ✓ | ✓ | × | - | ✓ | 77.8 | Low risk |
| Booij et al. 2015 | - | - | × | ✓ | ✓ | - | ✓ | ✓ | × | × | ✓ | 62.5 | High risk |
| Kaseva et al. 2019 | ✓ | ✓ | × | ✓ | ✓ | - | ✓ | ✓ | ✓ | ✓ | ✓ | 90 | Low risk |
| Criterion score % | 100 | 100 | 20 | 100 | 100 | - | 100 | 100 | 25 | 50 | 100 | | |

Note that the total risk of bias score was calculated by dividing the number of criteria met in one study by the total number of criteria (i.e., 10). Note that the criterion score is calculated by dividing the number of studies meeting one criterion by the total number of studies (i.e., 4). ✓: meet the methodological quality criterion; ×: not meet the methodological quality criterion; - Not applicable criterion.

**Table S9.** Risk assessment of included cross-sectional articles.

| Study | Item 1 | Item 2 | Item 3 | Item 4 | Item 5 | Item 6 | Item 7 | Item 8 | Quality score % | Risk category |
|---|---|---|---|---|---|---|---|---|---|---|
| Conley et al. 2020 | ✓ | × | ✓ | - | × | ✓ | ✓ | ✓ | 71.4 | High risk |
| Gorham et al. 2019 | ✓ | × | ✓ | - | × | ✓ | ✓ | ✓ | 71.4 | High risk |
| Babiss & Gangwisch 2009 | ✓ | ✓ | × | - | ✓ | ✓ | ✓ | ✓ | 85.7 | Low risk |
| Chae et al. 2017 | × | × | ✓ | - | ✓ | ✓ | ✓ | ✓ | 71.4 | High risk |
| Dishman et al. 2006 | × | × | ✓ | - | ✓ | ✓ | ✓ | ✓ | 71.4 | High risk |
| Gorham & Barch 2020 | ✓ | × | ✓ | - | × | ✓ | ✓ | ✓ | 71.4 | High risk |
| Herring et al. 2014 | × | × | ✓ | - | ✓ | ✓ | ✓ | ✓ | 71.4 | High risk |
| Joiner & Tickle 1998 | × | ✓ | ✓ | - | × | ✓ | ✓ | ✓ | 71.4 | High risk |
| Pickett et al. 2012 | ✓ | ✓ | ✓ | - | ✓ | ✓ | ✓ | ✓ | 100 | Low risk |
| Ansari et al. 2011 | × | ✓ | × | - | ✓ | ✓ | ✓ | ✓ | 71.4 | High risk |
| Kayani et al. 2021 | × | ✓ | ✓ | - | ✓ | ✓ | ✓ | ✓ | 85.7 | Low risk |
| Ryan et al. 2008 | × | × | ✓ | - | × | ✓ | ✓ | ✓ | 57.1 | High risk |
| Barham et al. 2021 | × | ✓ | × | - | × | ✓ | ✓ | ✓ | 57.1 | High risk |
| Criterion score % | 38.5 | 46.2 | 76.9 | - | 53.8 | 100 | 100 | 100 | | |

Note that the total risk of bias score was calculated by dividing the number of criteria met in one study by the total number of criteria (i.e., 10). Note that the criterion score is calculated by dividing the number of studies meeting one criterion by the total number of studies (i.e., 4). ✓: meet the methodological quality criterion; ×: not meet the methodological quality criterion; - Not applicable criterion

**Mechanisms linking physical activity with psychiatric symptoms across lifespan**

**Search strategies**

| Database searched | via | Years of coverage |
|---|---|---|
| Embase | Embase.com | 1971 - Present |
| Medline ALL | Ovid | 1946 - Present |
| Web of Science Core Collection* | Web of Knowledge | 1975 - Present |
| Cochrane Central Register of Controlled Trials | Wiley | 1992 - Present |
| PsycINFO | Ovid | 1806 - Present |

*Science Citation Index Expanded (1975-present) ; Social Sciences Citation Index (1975-present); Arts & Humanities Citation Index (1975-present) ; Conference Proceedings Citation Index- Science (1990-present); Conference Proceedings Citation Index- Social Science & Humanities (1990-present) ; Emerging Sources Citation Index (2015-present)

**Embase.com**

(exercise/exp OR 'physical activity'/de OR Climbing/exp OR running/exp OR walking/exp OR 'cycling'/de OR 'fighting'/de OR 'jogging'/de OR 'jumping'/de OR 'lifting effort'/de OR 'nordic walking'/de OR 'racewalking'/de OR 'stretching'/de OR 'swimming'/de OR 'weight bearing'/de OR 'weight lifting'/de OR sport/exp OR 'motor activity'/de OR 'psychomotor activity'/de OR 'physical education'/de OR locomotion/de OR swimming/de OR walking/exp OR (((physical* OR motor* OR psychomotor*) NEAR/3 (activit*)) OR exercise* OR sport* OR (physical NEAR/3 (education OR training)) OR locomotion* OR (Endurance NEAR/3 Training) OR walking OR running OR jogging OR yoga OR taichi OR tai-chi OR martial-art* OR qigong OR ((aerobic OR resistance OR physical*) NEAR/3 training)):ab,ti) AND (depression/exp OR 'anxiety disorder'/exp OR anxiety/de OR 'somatic symptom'/de OR 'attention deficit disorder'/de OR 'internalizing disorder'/de OR 'internalization (behavior)'/de OR 'externalizing disorder'/de OR 'externalization (behavior)'/de OR 'conduct disorder'/de OR 'rule breaking behavior'/de OR 'disruptive behavior'/exp OR (depressi* OR anxiet* OR (somatic* NEAR/3 symptom*) OR (attention NEAR/3 deficit NEAR/3 (disorder* OR hyperactiv* OR problem*)) OR (hyperactivit* NEAR/3 problem*) OR adhd OR internalizing OR internalization* OR internalising OR internalisation* OR externalizing OR externalization* OR externalising OR externalisation* OR (conduct NEAR/3 (disorder* OR problem*)) OR (rule* NEAR/3 breaking NEAR/3 behav*) OR ((disrupt* OR problem*) NEAR/3 behav*)):ab,ti) AND ('nuclear magnetic resonance imaging'/exp OR 'diffusion tensor imaging'/exp OR electroencephalography/exp OR electroencephalogram/exp OR 'gray matter'/exp OR 'white matter'/exp OR 'nervous system development'/exp OR 'nerve cell plasticity'/exp OR 'biological marker'/de OR marker/de OR hormone/exp OR peptide/exp OR protein/exp OR neurotransmitter/de OR opiate/de OR serotonin/de OR 'serotonin level'/exp OR noradrenalin/de OR dopamine/de OR monoamine/de OR endorphin/de OR neuroimmunology/de OR 'growth factor'/exp OR 'brain derived neurotrophic factor'/de OR 'neurotrophin gene'/de OR vasculotropin/de OR 'BOLD signal'/de OR volumetry/de OR 'brain function'/de OR 'functional connectivity'/de OR 'brain asymmetry'/de OR 'brain metabolism'/de OR 'brain structure'/de OR 'inflammatory marker'/de OR 'event related brain potential'/de OR 'emotional intelligence'/de OR 'social interaction'/de OR 'self esteem'/exp OR 'self control'/exp OR 'social connectedness'/exp OR 'self acceptance'/exp OR 'personal autonomy'/de OR belongingness/de OR confidence/de OR 'self confidence'/de OR 'self concept'/de OR (mri OR (magnetic NEAR/3 resonance) OR (diffusion NEAR/3 tensor NEAR/3 imag*) OR electroencephalogra* OR eeg OR ((grey OR gray OR white) NEXT/1 matter*) OR (nervous-system* NEAR/3 development*) OR neurogenesis* OR ((nerv*-cell* OR Neuron*) NEAR/3 plasticit*) OR hormon* OR peptide* OR protein* OR neurotransmitter* OR opiate* OR opioid* OR serotonin* OR noradrenalin* OR norepinephrin* OR dopamin* OR monoamin* OR endorphin* OR neuroimmunolo* OR neuro-immunolo* OR growth-factor* OR (brain-deriv* NEAR/3 neurotroph*-factor*) OR neurotrophin*-gene* OR IGF1 OR IGF-1 OR vasculotropin* OR VEGF OR BDNF OR VBM

OR BOLD-signal* OR (blood-oxygen NEAR/3 level* NEAR/3 dependent*) OR volumetr* OR (brain NEAR/3 (function* OR asymmetry* OR metabol* OR structure*)) OR (functional* NEAR/3 connectiv*) OR (event-related NEAR/3 brain-potential*) OR ((emotion* OR psycholog*) NEAR/3 (intelligen* OR adjustment* OR regulation*)) OR (social* NEAR/3 (interacti* OR connected*)) OR (self NEXT/1 (esteem OR control* OR acceptan* OR awareness* OR concept OR efficac*)) OR (psychologic* NEAR/3 need*) OR autonom* OR belongingness* OR (perceived NEAR/3 abilit*) OR (confidence NOT (confidence-interval*))):ab,ti) AND ('intervention study'/de OR 'clinical trial'/exp OR randomization/exp OR 'prospective study'/exp OR 'longitudinal study'/exp OR 'cohort analysis'/de OR 'follow up'/de OR 'cross-sectional study'/de OR 'major clinical study'/de OR (intervention* OR trial* OR random* OR rct OR prospective* OR longitudinal* OR cohort* OR follow-up* OR cross-section*):ab,ti) NOT ([animals]/lim NOT [humans]/lim) NOT (patient/exp/mj OR athlete/exp/mj OR 'breathing exercise'/mj OR 'degenerative disease'/exp/mj OR neoplasm/exp/mj OR 'cerebrovascular disease'/exp/mj OR 'cardiovascular disease'/exp/mj OR 'musculoskeletal disease'/exp/mj OR pain/exp/mj OR surgery/exp/mj OR survivor/exp/mj OR diabetes/exp/mj OR (patient* OR athlete* OR breathing OR disease* OR Alzheimer* OR Parkinson* OR osteoarthrit* OR cancer OR neoplas* OR stroke OR cva OR cerebrovascular OR cardiovascular OR musculoskeletal OR pain OR disorder* OR surger* OR transplant* OR survivor* OR diabet* OR injur*):ti) NOT [conference abstract]/lim NOT ('systematic review'/de OR 'meta analysis'/de OR 'case report'/de OR ((systematic* NEAR/3 review*) OR meta-analys* OR case-report*):Ab,ti)

**Medline ALL Ovid**

(Exercise/ OR Climbing/ OR Running/ OR Walking/ OR Bicycling/ OR Jogging/ OR Weight-Bearing/ OR Weight Lifting/ OR exp Sports/ OR Motor Activity/ OR "Physical Education and Training"/ OR Locomotion/ OR Swimming/ OR (((physical* OR motor* OR psychomotor*) ADJ3 (activit*)) OR exercise* OR sport* OR (physical ADJ3 (education OR training)) OR locomotion* OR (Endurance ADJ3 Training) OR walking OR running OR jogging OR yoga OR taichi OR tai-chi OR martial-art* OR qigong OR ((aerobic OR resistance OR physical*) ADJ3 training)).ab,ti.) AND (exp Depressive Disorder/ OR Depression/ OR exp Anxiety Disorders/ OR Anxiety/ OR Medically Unexplained Symptoms/ OR exp "Attention Deficit and Disruptive Behavior Disorders"/ OR Conduct Disorder/ OR Problem Behavior/ OR (depressi* OR anxiet* OR (somatic* ADJ3 symptom*) OR (attention ADJ3 deficit ADJ3 (disorder* OR hyperactiv* OR problem*)) OR (hyperactivit* ADJ3 problem*) OR adhd OR internalizing OR internalization* OR internalising OR internalisation* OR externalizing OR externalization* OR externalising OR externalisation* OR (conduct ADJ3 (disorder* OR problem*)) OR (rule* ADJ3 breaking ADJ3 behav*) OR ((disrupt* OR problem*) ADJ3 behav*)).ab,ti.) AND (exp Magnetic Resonance Imaging/ OR exp Diffusion Tensor Imaging/ OR Electroencephalography/ OR Gray Matter/ OR White Matter/ OR Biomarkers/ OR Hormones/ OR Peptides/ OR Proteins/ OR Neurotransmitter Agents/ OR Opiate Alkaloids/ OR Serotonin/ OR Norepinephrine/ OR Dopamine/ OR Endorphins/ OR Opioid Peptides/ OR Intercellular Signaling Peptides and Proteins/ OR Brain-Derived Neurotrophic Factor/ OR Vascular Endothelial Growth Factor A/ OR BOLD signal/ OR Emotional Intelligence/ OR Social Interaction/ OR Self-Control/ OR Personal Autonomy/ OR Self Concept/ OR (mri OR (magnetic ADJ3 resonance) OR (diffusion ADJ3 tensor ADJ3 imag*) OR electroencephalogra* OR eeg OR ((grey OR gray OR white) ADJ matter*) OR (nervous-system* ADJ3 development*) OR neurogenesis* OR ((nerv*-cell* OR Neuron*) ADJ3 plasticit*) OR hormon* OR peptide* OR protein* OR neurotransmitter* OR opiate* OR opioid* OR serotonin* OR noradrenalin* OR norepinephrin* OR dopamin* OR monoamin* OR endorphin* OR neuroimmunolo* OR neuro-immunolo* OR growth-factor* OR (brain-deriv* ADJ3 neurotroph*-factor*) OR neurotrophin*-gene* OR IGF1 OR IGF-1 OR vasculotropin* OR VEGF OR BDNF OR VBM OR BOLD-signal* OR (blood-oxygen ADJ3 level* ADJ3 dependent*) OR volumetr* OR (brain ADJ3 (function* OR asymmetry* OR metabol* OR structure*)) OR (functional* ADJ3 connectiv*) OR (event-related ADJ3 brain-potential*) OR ((emotion* OR psycholog*) ADJ3 (intelligen* OR adjustment* OR regulation*)) OR (social* ADJ3 (interacti* OR connected*)) OR (self ADJ (esteem OR control* OR acceptan* OR awareness* OR concept OR efficac*)) OR (psychologic* ADJ3 need*) OR autonom* OR belongingness* OR (perceived ADJ3 abilit*) OR (confidence NOT (confidence-interval*))).ab,ti.) AND (exp Clinical Trial/ OR Random Allocation/ OR exp Cohort Studies/ OR Cross-Sectional Studies/ OR (intervention* OR trial* OR random* OR rct OR prospective* OR longitudinal* OR cohort* OR follow-up* OR cross-section*).ab,ti.) NOT (exp animals/ NOT humans/) NOT (exp * Patients/ OR exp * Athletes/ OR * Breathing Exercises/ OR exp * Neoplasms/ OR exp * Cerebrovascular Disorders/ OR exp * Cardiovascular Diseases/ OR exp * Musculoskeletal Diseases/ OR *exp Pain/ OR exp * Surgical Procedures, Operative/ OR exp * Survivors/ OR *exp Diabetes Mellitus/ OR (patient* OR athlete* OR breathing OR disease* OR Alzheimer* OR Parkinson* OR osteoarthrit* OR cancer OR neoplas* OR stroke OR cva OR cerebrovascular OR cardiovascular OR musculoskeletal OR pain OR disorder* OR surger* OR transplant* OR survivor* OR diabet* OR injur*).ti.) NOT (Systematic Review/ OR Meta-Analysis/ OR case report/ OR ((systematic* ADJ3 review*) OR meta-analys* OR case-report*).ab,ti.)

**PSycINFO ALL Ovid**

(exp Exercise/ OR Physical Activity/ OR exp Sports/ OR (((physical* OR motor* OR psychomotor*) ADJ3 (activit*)) OR exercise* OR sport* OR (physical ADJ3 (education OR training)) OR locomotion* OR (Endurance ADJ3 Training) OR walking OR running OR jogging OR yoga OR taichi OR tai-chi OR martial-art* OR qigong OR ((aerobic OR resistance OR physical*) ADJ3 training)).ab,ti.) AND (exp Major Depression/ OR "Depression (Emotion)"/ OR exp Anxiety Disorders/ OR Anxiety/ OR exp Attention Deficit Disorder with Hyperactivity / OR Conduct Disorder/ OR Behavior Problems / OR (depressi* OR anxiet* OR (somatic* ADJ3 symptom*) OR (attention ADJ3 deficit ADJ3 (disorder* OR hyperactiv* OR problem*)) OR (hyperactivit* ADJ3 problem*) OR adhd OR internalizing OR internalization* OR internalising OR internalisation* OR externalizing OR externalization* OR externalising OR externalisation* OR (conduct ADJ3 (disorder* OR problem*)) OR (rule* ADJ3 breaking ADJ3 behav*) OR ((disrupt* OR problem*) ADJ3 behav*)).ab,ti.) AND (exp Magnetic Resonance Imaging/ OR exp Diffusion Tensor Imaging/ OR Electroencephalography/ OR Gray Matter/ OR White Matter/ OR Biological Markers / OR Hormones/ OR Peptides/ OR Proteins/ OR Neurotransmitters / OR Opiates / OR Serotonin/ OR Norepinephrine/ OR Dopamine/ OR Endorphins/ OR Brain Derived Neurotrophic Factor / OR Emotional Intelligence/ OR Social Interaction/ OR Self-Control/ OR Autonomy/ OR Self-Concept/ OR (mri OR (magnetic ADJ3 resonance) OR (diffusion ADJ3 tensor ADJ3 imag*) OR electroencephalogra* OR eeg OR ((grey OR gray OR white) ADJ matter*) OR (nervous-system* ADJ3 development*) OR neurogenesis* OR ((nerv*-cell* OR Neuron*) ADJ3 plasticit*) OR hormon* OR peptide* OR protein* OR neurotransmitter* OR opiate* OR opioid* OR serotonin* OR noradrenalin* OR norepinephrin* OR dopamin* OR monoamin* OR endorphin* OR neuroimmunolo* OR neuro-immunolo* OR growth-factor* OR (brain-deriv* ADJ3 neurotroph*-factor*) OR neurotrophin*-gene* OR IGF1 OR IGF-1 OR vasculotropin* OR VEGF OR BDNF OR VBM OR BOLD-signal* OR (blood-oxygen ADJ3 level* ADJ3 dependent*) OR volumetr* OR (brain ADJ3 (function* OR asymmetry* OR metabol* OR structure*)) OR (functional* ADJ3 connectiv*) OR (event-related ADJ3 brain-potential*) OR ((emotion* OR psycholog*) ADJ3 (intelligen* OR adjustment* OR regulation*)) OR (social* ADJ3 (interacti* OR connected*)) OR (self ADJ (esteem OR control* OR acceptan* OR awareness* OR concept OR efficac*)) OR (psychologic* ADJ3 need*) OR autonom* OR belongingness* OR (perceived ADJ3 abilit*) OR (confidence NOT (confidence-interval*))).ab,ti.) AND (exp Clinical Trials / OR exp Cohort Analysis / OR (intervention* OR trial* OR random* OR rct OR prospective* OR longitudinal* OR cohort* OR follow-up* OR cross-section*).ab,ti.) NOT (exp animals/ NOT humans/) NOT (exp * Patients/ OR exp * Athletes/ OR exp * Neoplasms/ OR exp * Cerebrovascular Disorders/ OR exp * Cardiovascular Disorders / OR exp * Musculoskeletal Disorders / OR *exp Pain/ OR exp * Surgery / OR exp * Survivors/ OR *exp Diabetes Mellitus/ OR (patient* OR athlete* OR breathing OR disease* OR Alzheimer* OR Parkinson* OR osteoarthrit* OR cancer OR neoplas* OR stroke OR cva OR cerebrovascular OR cardiovascular OR musculoskeletal OR pain OR disorder* OR surger* OR transplant* OR survivor* OR diabet* OR injur*).ti.) NOT ("Systematic Review"/ OR Meta Analysis/ OR case report/ OR ((systematic* ADJ3 review*) OR meta-analys* OR case-report*).ab,ti.)

**Cochrane CENTRAL register of Trials**

((((physical* OR motor* OR psychomotor*) NEAR/3 (activit*)) OR exercise* OR sport* OR (physical NEAR/3 (education OR training)) OR locomotion* OR (Endurance NEAR/3 Training) OR walking OR running OR jogging OR yoga OR taichi OR tai NEXT chi OR martial NEXT art* OR qigong OR ((aerobic OR resistance OR physical*) NEAR/3 training)):ab,ti) AND ((depressi* OR anxiet* OR (somatic* NEAR/3 symptom*) OR (attention NEAR/3 deficit NEAR/3 (disorder* OR hyperactiv* OR problem*)) OR (hyperactivit* NEAR/3 problem*) OR adhd OR internalizing OR internalization* OR internalising OR internalisation* OR externalizing OR externalization* OR externalising OR externalisation* OR (conduct NEAR/3 (disorder* OR problem*)) OR (rule* NEAR/3 breaking NEAR/3 behav*) OR ((disrupt* OR problem*) NEAR/3 behav*)):ab,ti) AND ((mri OR (magnetic NEAR/3 resonance) OR (diffusion NEAR/3 tensor NEAR/3 imag*) OR electroencephalogra* OR eeg OR ((grey OR gray OR white) NEXT/1 matter*) OR (nervous NEXT system* NEAR/3 development*) OR neurogenesis* OR ((nerv* NEXT cell* OR Neuron*) NEAR/3 plasticit*) OR hormon* OR peptide* OR protein* OR neurotransmitter* OR opiate* OR opioid* OR serotonin* OR noradrenalin* OR norepinephrin* OR dopamin* OR monoamin* OR endorphin* OR neuroimmunolo* OR neuro NEXT immunolo* OR growth NEXT factor* OR (brain NEXT deriv* NEAR/3 neurotroph* NEXT factor*) OR neurotrophin* NEXT gene* OR IGF1 OR IGF NEXT 1 OR vasculotropin* OR VEGF OR BDNF OR VBM OR BOLD NEXT signal* OR (blood NEXT oxygen NEAR/3 level* NEAR/3 dependent*) OR volumetr* OR (brain NEAR/3 (function* OR asymmetry* OR metabol* OR structure*)) OR (functional* NEAR/3 connectiv*) OR (event NEXT related NEAR/3 brain NEXT potential*) OR ((emotion* OR psycholog*) NEAR/3 (intelligen* OR adjustment* OR regulation*)) OR (social* NEAR/3 (interacti* OR connected*)) OR (self NEXT/1 (esteem OR control* OR acceptan* OR awareness* OR concept OR efficac*)) OR (psychologic* NEAR/3 need*) OR autonom* OR belongingness* OR (perceived NEAR/3 abilit*) OR (confidence NOT (confidence NEXT interval*))):ab,ti) NOT (patient* OR athlete* OR breathing OR disease* OR Alzheimer* OR Parkinson* OR osteoarthrit* OR cancer OR neoplas* OR stroke OR cva OR cerebrovascular OR cardiovascular OR musculoskeletal OR pain OR disorder* OR surger* OR transplant* OR survivor* OR diabet* OR injur*):ti

**Web of Science Core Collection**

TS=(((((physical* OR motor* OR psychomotor*) NEAR/2 (activit*)) OR exercise* OR sport* OR (physical NEAR/2 (education OR training)) OR locomotion* OR (Endurance NEAR/2 Training) OR walking OR running OR jogging OR yoga OR taichi OR tai-chi OR martial-art* OR qigong OR ((aerobic OR resistance OR physical*) NEAR/2 training))) AND ((depressi* OR anxiet* OR (somatic* NEAR/2 symptom*) OR (attention NEAR/2 deficit NEAR/2 (disorder* OR hyperactiv* OR problem*)) OR (hyperactivit* NEAR/2 problem*) OR adhd OR internalizing OR internalization* OR internalising OR internalisation* OR externalizing OR externalization* OR externalising OR externalisation* OR (conduct NEAR/2 (disorder* OR problem*)) OR (rule* NEAR/2 breaking NEAR/2 behav*) OR ((disrupt* OR problem*) NEAR/2 behav*))) AND ((mri OR (magnetic NEAR/2 resonance) OR (diffusion NEAR/2 tensor NEAR/2 imag*) OR electroencephalogra* OR eeg OR ((grey OR gray OR white) NEAR/1 matter*) OR (nervous-system* NEAR/2 development*) OR neurogenesis* OR ((nerv*-cell* OR Neuron*) NEAR/2 plasticit*) OR hormon* OR peptide* OR protein* OR neurotransmitter* OR opiate* OR opioid* OR serotonin* OR noradrenalin* OR norepinephrin* OR dopamin* OR monoamin* OR endorphin* OR neuroimmunolo* OR neuro-immunolo* OR growth-factor* OR (brain-deriv* NEAR/2 neurotroph*-factor*) OR neurotrophin*-gene* OR IGF1 OR IGF-1 OR vasculotropin* OR VEGF OR BDNF OR VBM OR BOLD-signal* OR (blood-oxygen NEAR/2 level* NEAR/2 dependent*) OR volumetr* OR (brain NEAR/2 (function* OR asymmetry* OR metabol* OR structure*)) OR (functional* NEAR/2 connectiv*) OR (event-related NEAR/2 brain-potential*) OR ((emotion* OR psycholog*) NEAR/2 (intelligen* OR adjustment* OR regulation*)) OR (social* NEAR/2 (interacti* OR connected*)) OR (self NEAR/1 (esteem OR control* OR acceptan* OR awareness* OR concept OR efficac*)) OR (psychologic* NEAR/2 need*)

OR autonom* OR belongingness* OR (perceived NEAR/2 abilit*) OR (confidence NOT (confidence-interval*)))) AND ((intervention* OR trial* OR random* OR rct OR prospective* OR longitudinal* OR cohort* OR follow-up* OR cross-section*))) NOT TI=(patient* OR athlete* OR breathing OR disease* OR Alzheimer* OR Parkinson* OR osteoarthrit* OR cancer OR neoplas* OR stroke OR cva OR cerebrovascular OR cardiovascular OR musculoskeletal OR pain OR disorder* OR surger* OR transplant* OR survivor* OR diabet* OR injur* OR (systematic* NEAR/3 review*) OR meta-analys* OR case-report*) AND DT=(article) AND LA=(english)
